# Peripheral and lung resident T cell responses against SARS-CoV-2

**DOI:** 10.1101/2020.12.02.20238907

**Authors:** Judith Grau-Expósito, Nerea Sánchez-Gaona, Núria Massana, Marina Suppi, Antonio Astorga-Gamaza, David Perea, Joel Rosado, Anna Falcó, Cristina Kirkegaard, Ariadna Torrella, Bibiana Planas, Jordi Navarro, Paula Suanzes, Daniel Alvarez-de la Sierra, Alfonso Ayora, Irene Sansano, Juliana Esperalba, Cristina Andrés, Andrés Antón, Santiago Ramón y Cajal, Benito Almirante, Ricardo Pujol-Borrell, Vicenç Falcó, Joaquín Burgos, María J. Buzón, Meritxell Genescà

**Affiliations:** Infectious Diseases Department, Vall d’Hebron Institut de Recerca (VHIR), Vall d’Hebron Hospital Universitari, Vall d’Hebron Barcelona Hospital Campus, Passeig Vall d’Hebron 119-129, 08035 Barcelona, Spain; Thoracic Surgery and Lung Transplantation Department, Vall d’Hebron Institut de Recerca (VHIR), Vall d’Hebron Hospital Universitari, Vall d’Hebron Barcelona Hospital Campus, Passeig Vall d’Hebron 119-129, 08035 Barcelona, Spain; Diagnostic Immunology Group, Vall d’Hebron Institut de Recerca (VHIR), Vall d’Hebron Hospital Universitari, Vall d’Hebron Barcelona Hospital Campus, Passeig Vall d’Hebron 119-129, 08035 Barcelona, Spain; Occupational Risk Prevention Unit, Vall d’Hebron Hospital Universitari, Vall d’Hebron Barcelona Hospital Campus, Passeig Vall d’Hebron 119-129, 08035 Barcelona, Spain; Pathology Department, Vall d’Hebron Hospital Universitari, Vall d’Hebron Barcelona Hospital Campus, Passeig Vall d’Hebron 119-129, 08035 Barcelona, Departament de Ciències morfològiques, Universitat Autònoma de Barcelona, Universitat Autònoma de Barcelona, 08193 Bellaterra, Spain; Respiratory Viruses Unit, Microbiology Department, Vall d’Hebron Institut de Recerca (VHIR), Vall d’Hebron Hospital Universitari, Vall d’Hebron Barcelona Hospital Campus, Passeig Vall d’Hebron 119-129, 08035 Barcelona, Spain; FOCIS Center of Excellence

## Abstract

Considering that SARS-CoV-2 interacts with the host at the respiratory tract mucosal interface, T cells strategically placed within these surfaces, namely resident memory T cells, will be essential to limit viral spread and disease. Importantly, these cells are mostly non-recirculating, which reduces the window of opportunity to examine circulating lymphocytes in blood as they home to the lung parenchyma. Here, we demonstrate that viral specific T cells can migrate and establish in the lung as resident memory T cells remaining detectable up to 10 months after initial infection. Moreover, focusing on the acute phase of the infection, we identified virus-specific T cell responses in blood with functional, migratory and apoptotic patterns modulated by viral proteins and associated with clinical outcome. Our study highlights IL-10 secretion by virus-specific T cells associated to a better outcome and the persistence of resident memory T cells as key players for future protection against SARS-CoV-2 infection.

## INTRODUCTION

We are currently facing a global health emergency, the COVID-19 pandemic. While a great effort is focused on vaccine development, many questions remain unanswered that are necessary to properly manage patients and inform vaccine assessment. To this end, identifying the development of a protective immune response after natural infection and characterizing the correlates of protection would greatly inform on the best strategy to stimulate a protective response by immunization. Moreover, identifying specific immunological parameters capable of predicting disease control (i.e. no hospitalization) could provide new biomarkers to support medical decisions for patients. Most efforts to measure or induce immunity rely on neutralizing antibodies which can certainly limit infection; however, antibody detection is not only inconsistent in infected or convalescent patients^1, 2, 3, 4^ but may also wane with time as shown for other coronaviruses^5, 6^, although their stability may also depend on the antigen targeted ^7^.

Virus-specific T cells against SARS-CoV-2 have been shown to develop against coronaviruses^, 8, 9, 10, 11, 12, 13, 14^ and specific memory T cells persisted in SARS-recovered patients for up to 6 years post-infection^5^. Thus, T cells may potentially provide long-term immunity as demonstrated for other viral infections such as SARS or influenza^15, 16, 17, 18^. In this sense, mouse models of SARS-CoV-1 infection demonstrated that both CD8^+^T and CD4^+^T cells are critical for viral clearance^15, 17^. Considering that SARS-CoV-2 interacts with the host at the respiratory tract mucosal interface, T cells strategically positioned within these surfaces, may be essential to limiting infection. A key role for resident memory T cells (T_RM_) in protection against pathogen challenge has been established for many tissues, including the lung^5, 17, 19, 20, 21, 22^. These cells, which are strategically located both in the lung airways and in lung interstitial tissue, include CD4^+^ and CD8^+^T cells designed to limit re-infections locally. In the context of respiratory infection models such as influenza, CD8^+^T_RM_ have shown to confer cross-protection against different strains^22^, and both influenza-specific CD4^+^ and CD8^+^T_RM_ have been identified^16, 23^. Importantly, optimal protection against SARS-CoV-infected mice was conferred by airway memory CD4^+^T cells secreting both, pro-inflammatory interferon gamma (IFNg) and anti-inflammatory interleukin(IL)-10 molecules^17^. Thus, a broader spectrum of T helper (Th) profiles should be included when addressing virus-specific T cells. Further, recruitment of these cells from circulation may depend on the expression of molecules such as chemokine C-X-C motif receptor 3 (CXCR3), which, besides mediating chemotaxis toward inflamed tissue of a biased Th1 profile, appears critical for the recruitment of pulmonary T cells that control infection^18, 24, 25^. While such recruitment could partially contribute to the decrease of circulating lymphocytes and thus favor tissue infiltration, many other factors may explain the observed lymphopenia associated with COVID-19 disease severity^26, 27, 28, 29^. In this regard, increased susceptibility of both specific and bystander CD4^+^ and CD8^+^T lymphocytes to apoptotic cell death has been observed in other viral infections^30, 31, 32^, which could be linked to increased glycolysis^33, 34^.

Here, we address several key questions related to early control of SARS-CoV-2 infection mediated by cellular immunity and long-term protection: 1) the functional profile of antigen-specific T cells associated with disease control; 2) if apoptosis is involved in disease severity; 3) if antigen-specific T cell responses have the potential to migrate to the lung and eventually become T_RM_ cells. To this end, we performed detailed phenotypic and functional analyses in clinically-defined groups of patients recruited during the first wave of SARS-CoV-2 infection, including the assessment of T_RM_ in lung of convalescent patients. Informing on the immunological parameters associated with disease control and patient prognosis will aid vaccine development and monitoring of vaccinated individuals towards prediction of immune control.

## RESULTS

### Cohort characteristics

Patients were recruited during the first pandemic wave of SARS-CoV-2 (spring 2020). A total of 46 patients were included, in which 14 individuals were symptomatic non-hospitalized cases, 20 individuals corresponded to mild-hospitalized cases and 12 to severe-hospitalized cases. Only one patient from the severe group corresponded to a fatal case. Samples were obtained between 7-16 days after symptom onset and no differences between groups were detected (Figure S1A). Table S1 shows a summary of the participant characteristics and baseline determinations, in which significant differences are evidenced between the three groups. As previously reported^26, 28, 29, 35, 36^, age, lymphopenia, and biochemical parameters such as D-dimer, IL-6, and ferritin were associated with disease severity. Quantification of the viral load between days 5 and 15 after symptom onset is also reported; however, values between the groups were not statistically significant. Some of the clinical parameters used to stratify mild and severe hospitalized cases are shown in Figure S1B: days to discharge since symptoms onset (p<0.0001), baseline IL-6 (p=0.008) and the percentage of oxyhemoglobin saturation in arterial blood /fraction of inspired oxygen (SAFI) ratio at baseline (p=0.0052) and after 48h (p=0.0002). These parameters were used to address associations between immunological parameters and disease severity. In some analyses, 12 control individuals sampled before the COVID-19 pandemic were studied in parallel.

To determine if whole plasma cytokine levels in our groups of COVID-19 patients were similar to previously defined patterns reported before^29, 37, 38, 39, 40^, we analyzed cytokine plasma levels by the ELLA® microfluidics platform in the same samples. Levels of IL-1ra, IL-2, IL-6, IL- 10, IL-15, CXCL10 (IP-10), IFNg, granzyme B and TNFa were elevated in the plasma of hospitalized groups compared to non-hospitalized patients, with higher levels associated with disease severity (Figure S1C). Of note, the deceased patient from the severe cohort (circled in green) had very high levels of some of the molecules associated with severity and fatality prediction, namely IL-6 and IL-1ra^38, 40^. Further, CXCL10 was the most significant predictor of hospitalization during acute infection, potentially related to impaired T cell responses as suggested^13^. CCL2, also referred to as monocyte chemoattractant protein 1, was significantly higher in the severe-hospitalized group compared to the non-hospitalized patients, while IL-4, IL- 7, IL-13, IL-17A, GM-CSF were similar among all three groups of patients (Figure S1C). Strikingly, the levels of IL-12p70 were higher in the plasma of the non-hospitalized compared to the mild COVID-19 group, and the deceased patient had the second lowest level of IL-12p70 of the severe group (0.093pg/mL; Figure S1B).

### Functional patterns associated to acute infection are defined by disease severity and targeted antigen

The functional capabilities of specific CD4+ and CD8^+^T cells against SARS-CoV-2 were measured by intracellular cytokine staining in samples ranging from 7 to 16 days-post-symptom onset (mean of 12 days) from all three groups. For that, we stimulated peripheral blood mononuclear cells (PBMC) with overlapping membrane (M), nucleocapsid (N) and spike (S) peptide sets and determined the expression of IFNg, IL-4, and IL-10, along with the degranulation marker CD107a in CD4^+^ and CD8^+^T cells (Figure S2A and 2B). For each function we calculated the net response for each peptide set (background subtracted) and compared these antigen-specific T cell responses among all three groups (Figure 1A). This way, differences on the frequency of IFNg- secreting antigen-specific T cells were significantly higher among the hospitalized groups compared to the outpatients (in CD4^+^T cells: p=0.020 for M and S peptides in the mild group; p=0.004 for M, p=0.011 for N and p=0.007 for S peptides in the severe group; Figure 1A). Of note, while non-hospitalized patients did not show a significant increase in the production of IFNg as a group, some individuals did show an increase in their response (>0.02% after background subtraction) representing 43% of responders, which was lower than the 80% and 92% of responders observed in mild and severe hospitalized groups. In contrast degranulation, measured by CD107a expression, was less detected in general and significance among the groups was only reached in response to M peptides in severe patients compared to non-hospitalized patients (p=0.036) (Figure 1A). We also calculated double positive IFNg/CD107a CD8^+^T cells, as a surrogate of cytotoxic polyfunctional cells, since double positive cells could be detected in some patients, as exemplified in Figure S2. Interestingly, while the frequency of double positive CD107a^+^IFNg^+^CD8^+^T cells responding to the M peptides positively correlated with viral load, the same subset specific for N peptides inversely correlated with baseline levels of IL-6 within the hospitalized cohort (Figure S3A).

**Figure 1.**
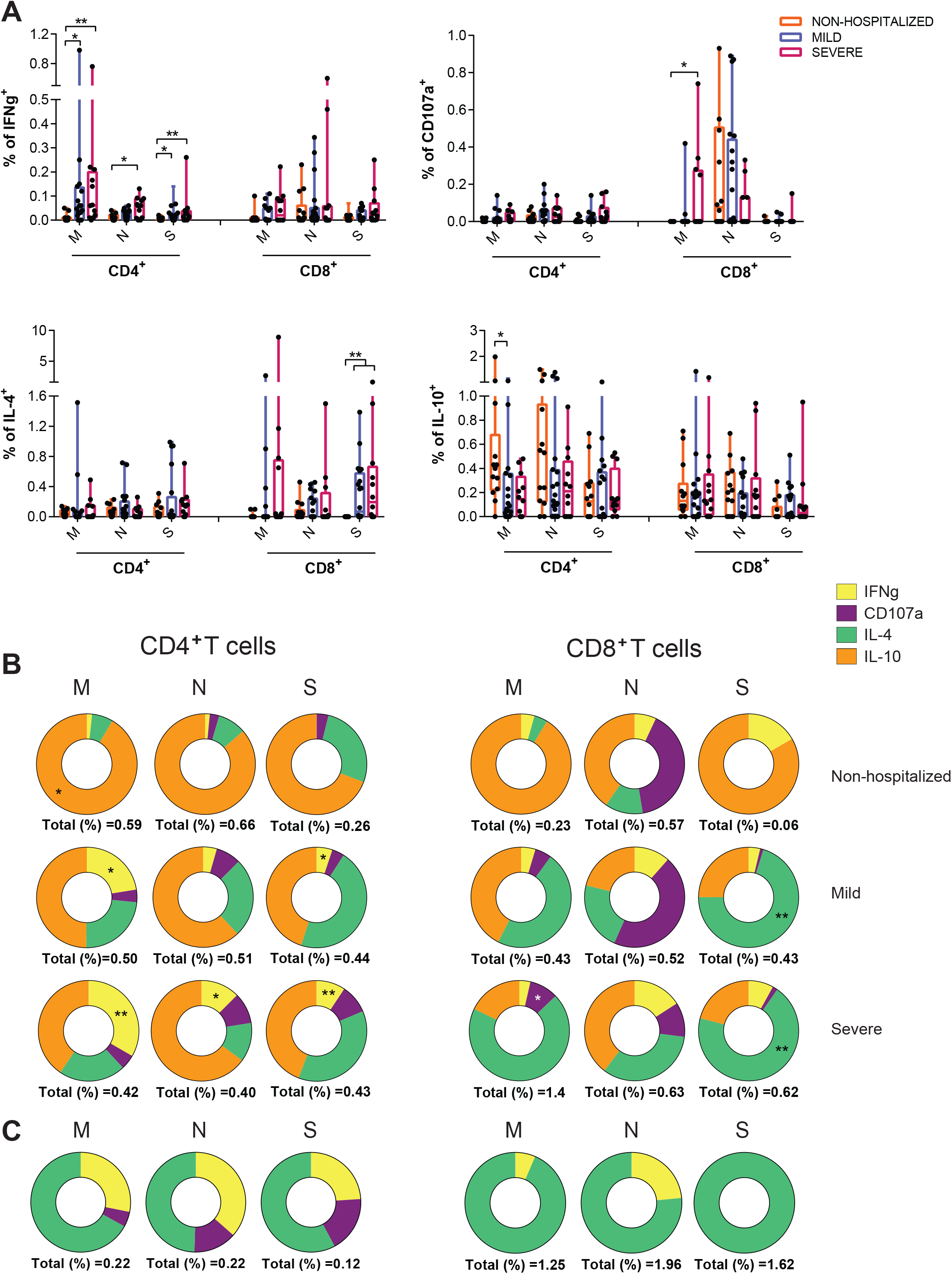
Functional characteristics of SARS-CoV-2-specific T cells by group and viral protein. **(A)** Comparison of the net frequency (background subtracted) of IFNg, CD107a, IL-4 and IL-10 expression in SARS-CoV-2-specific CD4^+^ and CD8^+^T cells in response to viral proteins (membrane (M), nucleocapsid (N) and spike (S)) between study groups (non-hospitalized n=14 in orange; mild n=20 in purple and severe n=12 in pink). Statistical comparisons were performed using Kruskal-Wallis rank-sum test with Dunn’s multiple comparison test. *p<0.05, **p<0.01. **(B)** Donut charts summarizing the contribution of each function to the overall CD4^+^ and CD8^+^ specific T cell response by targeted viral protein and individual group of patients. Data represents the mean value of the net frequency of each function indicated by color code considering all patients, responders and non-responders. Total response value (%) is shown under each pie chart and represents the cohort average of the overall net frequency considering all individuals and adding up all functions (non-hospitalized n=14; mild n=20 and severe n=12). **(C)** Donut charts summarizing the distribution of individual functions among specific-CD4^+^ and CD8^+^T cells to either the M, N or S protein from the only fatal case within the severe COVID-19 group.

Assessment of two other functions, IL-4 and IL-10, demonstrated major dominance of these responses based on the cohort and the viral target. A general induction of an IL-4-specific CD8^+^T cell response was observed in response to the viral spike in hospitalized patients compared to the non-hospitalized individuals (p=0.004 and p=0.003 for mild and severe patients, respectively; Figure 1A). Of note, higher levels of spontaneous secretion of IL-4 (in unstimulated conditions) were observed in hospitalized patients, which essentially correlated with the number of days since symptoms onset to discharge and baseline IL-6 levels (Figure S3B). Moreover, SARS-CoV-2 viral load positively correlated with the overall capacity of CD4^+^T cells to secrete IL- 4 in response to TCR independent unspecific activation with PMA/ionomycin (PMA/Io) (Figure S3C). In contrast, the expression of IL-10, a prototypical regulatory cytokine, was significantly increased in CD4^+^T cells from non-hospitalized patients after stimulation with M peptides when compared to the mild COVID-19 group (p=0.035; Figure 1A).

Correlations between the net frequency of a given function and clinical parameters were consistent with more CD4^+^T cells secreting IFNg and more CD8^+^T cells secreting IL-4 in response to M and S peptides associated with disease severity (Table S2). Even the total CD4^+^ or CD8^+^T cell IFNg response and the total IL-4 secretion by CD8^+^T cells against any of the three viral proteins (all peptides) correlated with more days at the hospital for IFNg or with other clinical parameters for IL-4 (Table S2). Further, antigen-specific CD4^+^T cells degranulating in response to all viral peptides and, in the case of CD8^+^T cells, in response to M peptides also correlated with higher levels of IL-6 (Table S2). In contrast, the percentage of M-specific CD4^+^T cells secreting IL-10 correlated with better prognosis in all clinical parameters (Table S2) and for N-specific positively with better oxygenation at 48h (Table S2).

Actually, when the overall response, including all functions, was represented as donut charts displaying the mean frequency of responses including all individuals (responders and non- responders), differences among groups in response to each peptide set were emphasized (Figure 1B). This was, M peptides were shown to mostly stimulate IL-10 secretion in non-hospitalized patients, while in hospitalized cases, increasing amounts of IFNg for CD4^+^T cells and of IL-4 and degranulation for CD8^+^T cell were observed (Figure 1B). In addition, N peptides induced higher frequencies of antigen-specific CD8^+^T cells degranulating in mild and non-hospitalized cases, while S peptides stimulated IL-4 secretion mainly in the hospitalized groups (Figure 1B). Overall, our analyses indicated, on one hand, group-based differences, where a dominance of IL-4 and IFNg SARS-CoV-2-specific responses were associated with disease severity and of IL-10 to minor disease; on the other hand, we observed targeted protein based-differences, where M and N peptides induced a Th1 profile exemplified by IFNg in CD4^+^T cells and degranulation (CD107a) in CD8^+^T cells, respectively, and S peptides induced a biased Th2 profile exemplified by IL-4. This pattern was shown in an exaggerated manner in the deceased patient, in which IL-10 responses were absent and IL-4 together with some IFNg dominated antigen-specific responses (Figure 1C).

### Expression patterns of chemokine receptors associated to SARS-CoV-2-infected patients

Next, we aimed to determine if part of the specific T cell response was potentially migrating towards the infected tissues by assessing the proportion of CCR7 and CXCR3 expression within the same analyses. In peripheral blood, CCR7 distinguishes T cells homing to lymph node (LN) when expressed, or effector memory (EM) T cell subsets migrating to tissues when absent^41^, while CXCR3 may help define antiviral T cells infiltrating inflamed tissues, including the lung parenchyma^25^. CD4^+^T cells showed only two evident subsets in most patients based on CCR7 expression, since CXCR3 was homogeneously dimly expressed in these two subsets (Figure S2A) and no differences between the different study groups were observed (Figure 2A). In contrast, CD8^+^T cells presented five subsets based on these chemokine receptors (Figure S2A and 2C), and significant differences among the groups were observed (Figure 2B). Non- hospitalized patients showed increased frequencies of CCR7^h^CXCR3^d^CD8^+^T cells, while severe patients presented increased frequencies of EM CCR7^-^CXCR3^+^T cells (p=0.0012 and p=0.0034 respectively, Figure 2B and 2C). In fact, the accumulation of CCR7^h^CXCR3^d^CD8^+^T cells indicated good prognosis and negatively correlated with the number of days to discharge since symptoms onset and with IL-6 levels at hospital entry (Figure 2D), while the frequency of EM CXCR3^+^ CD8^+^T cells significantly correlated with disease severity parameters (Figure 2E).

**Figure 2.**
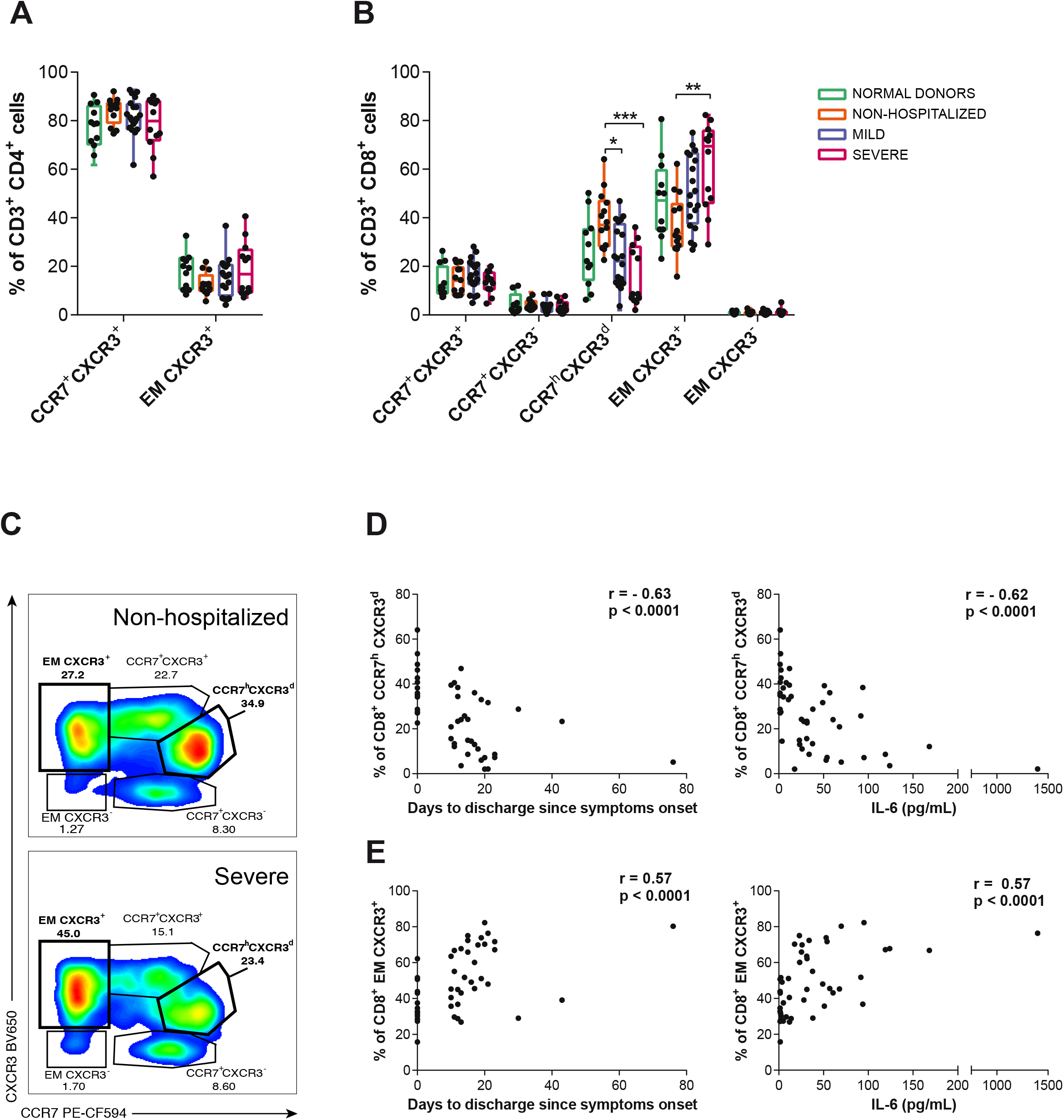
T cell migratory patterns during acute SARS-CoV-2 infection. **(A and B)** The frequency of various T cell subsets defined by CCR7 and CXCR3 within CD4^+^ **(A)** and CD8^+^T cells **(B)**. Each dot represents one patient of a specific cohort, indicated by color code (normal donors n=12; non-hospitalized n=14; mild n=20 and severe n=12). Data are shown as median and min to max range. Statistical comparisons were performed using Kruskal-Wallis rank- sum test with Dunn’s multiple comparison test. *p<0.05, **p<0.01, ***p<0.001. **(C)** Representative flow cytometry plots gating the different CD8^+^T cell subsets in a non-hospitalized (top) and a severe patient (bottom). **(D and E)** Correlations between days to discharge since symptoms onset or IL-6 baseline levels and the frequency of CD8^+^ CCR7^h^CXCR3^d^ **(D)** and CD8^+^ EM CXCR3^+^ **(E)** subpopulations. Spearman rank correlation (n=46).

We then focused on the distribution of the net antigen-response for each peptide and cohort among these CCR7/CXCR3 subsets. Overall, antigen-specific CD4^+^T cells showed a distinct pattern based on the function assessed: IFNg, degranulation (CD107a) and IL-4 were significantly associated to EM CXCR3^+^ CD4^+^T cells across the different groups and proteins, while IL-10 was associated to the LN-homing fraction (CCR7^+^CXCR3^+^) in response to M and N protein peptides in the non-hospitalized group (Figure 3A). Of note, in general, most individuals in the hospitalized groups also showed this trend for the IL-10 response, although statistical significance was not reached as a group. Moreover, several subsets out of these antigen-specific CD4^+^T cells, mostly the ones secreting IFNg or IL-4, correlated with worse prognosis in the clinical parameters assessed before, and some examples are shown in Figure 3B-3E. In general, stronger associations were observed within the CCR7^+^CXCR3^+^ subset, which correlated with severity, except if this subset was secreting IL-10 against M peptides (Figure 3F). Moreover, SARS-CoV- 2 viral load was negatively associated with the overall capacity of EM CXCR3^+^ CD4^+^T cells to secrete IL-10 in response to TCR independent unspecific activation with PMA/Io (Figure 3G).

**Figure 3.**
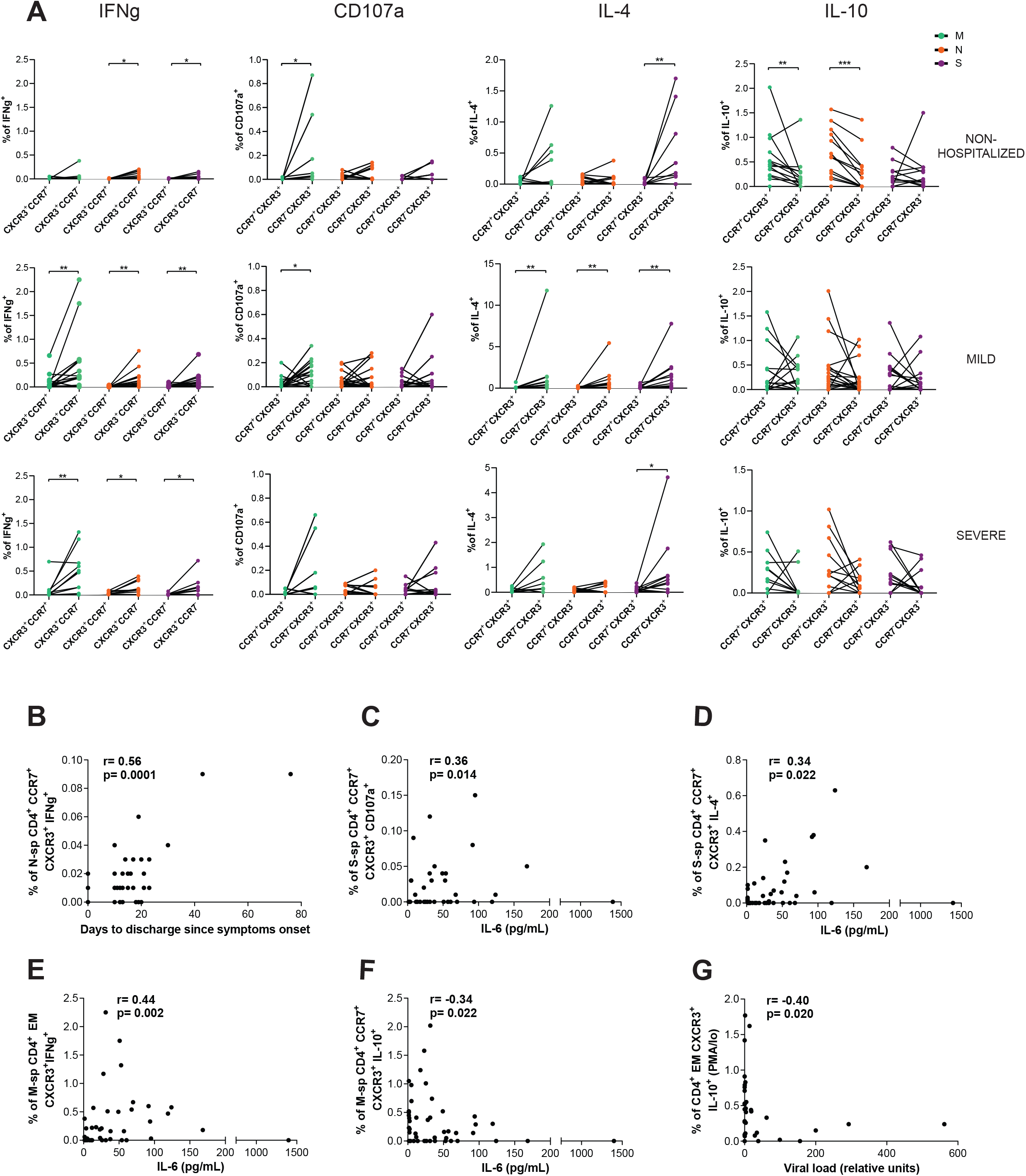
Migratory patterns of SARS-CoV-2-specific CD4^+^T cells expressing IFNg, CD107a, IL-4 or IL-10 by group and viral protein. **(A)** Net frequency of IFNg, CD107a, IL-4 and IL-10 expression in SARS-CoV-2-specific CD4^+^ T cells based on CXCR3^+^CCR7^+^ and CXCR3^+^CCR7^-^ subsets for each individual patient (non- hospitalized n=14; mild n=20 and severe n=12). Viral proteins are shown in color green (membrane protein, M), orange (nucleocapsid protein, N) and purple (spike protein, S). Dots connected by the same line represent the same individual. Statistical comparisons were performed using non-parametric Wilcoxon matched-pairs signed rank test to compare the two groups (CXCR3+CCR7^+^ vs. CXCR3^+^CCR7^-^). *p<0.05, **p<0.01, ***p<0.001. **(B-D)** Correlation between the days to discharge since symptoms onset or IL-6 and the frequency of nucleocapsid or spike-specific CD4^+^ CXCR3^+^CCR7^+^ expressing IFNg **(B)**, CD107a **(C)** or IL-4 **(D). (E and F)** Correlation between IL-6 and the frequency of membrane-specific CD4^+^ CXCR3^+^CCR7^-/+^ expressing IFNg **(E)** or IL-10 **(F). (G)** Correlation between the viral load and the frequency of CD4^+^ EM CXCR3^+^ expressing IL-10 after PMA/Ionomycin stimulation. Spearman rank correlation (n=46 for all correlations except for viral load **(G)** which corresponds to n=33).

In the analyses of the proportion of antigen-specific CD8^+^T cells in each CCR7/CXCR3 subset, we did not consider the EM CXCR3^-^ subset, which represented <1% in most patients (Figure 2B). As expected, IFNg antigen-specific CD8^+^T cells were more frequent among the CXCR3^+^ subsets, with some individual exception, such as N-specific CCR7^+^CXCR3^-^ T cells within the non-hospitalized group (Figure S4A). Further, IFNg secreting CCR7^h^CXCR3^d^CD8^+^T cells in response to N peptides correlated negatively with days to hospital discharge (Figure S4B). Degranulation, which was enhanced also after N stimulation, was unexpectedly detected in all CD8^+^ CCR7/CXCR3 subpopulations (Figure S5A). Though only in the LN-homing CCR7^+^CXCR3^+^CD8^+^T cell subset in response to S peptides, degranulation was associated with higher viral load (Figure S5B). With respect to IL-4 secreting antigen-specific CD8^+^T cells, in general these responses were more frequent in CCR7^+^ LN-homing subsets, and as mentioned before, they increased with disease severity (Figure S6A). Consequently, the frequency of IL-4 detected in response to M or S peptides in several of these fractions correlated with disease severity (Figure S6B). Remarkably, SARS-CoV-2-specific CD8^+^T cells secreting IL-10 were strongly represented among the CCR7^h^CXCR3^d^ subset, reaching statistical significance in response to any of the viral proteins within the mild disease cohort and in response to N peptides in non-hospitalized patients, but not in the hospitalized group with severe disease (Figure 4A). Finally, we detected two additional correlations within the CD8^+^T cell compartment that were of interest: overall antigen-specific EM CXCR3^+^ CD8^+^T cells correlated with higher viral loads if responding to M peptides (Figure 4B), while the same subset responding to N peptides negatively correlated with IL-6 (Figure 4C). All together, these results demonstrate individual migratory patterns associated with a given function: whilst most of the functions assessed here were associated with lung homing subsets (CCR7^-^), IL-10-specific T cells expressed high levels of CCR7. In fact, a strong association towards better disease prognosis was established for an increased proportion of CCR7^h^CXCR3^d^ CD8^+^T cells, which represented a major constant source of IL-10 in CD8^+^T cells. Further, antigenic stimulation could be driving CCR7^-^ effector immune responses towards the lung, yet under uncontrolled disease progression, such effector functions seemed to increase in LN-homing CCR7^+^ subsets.

**Figure 4.**
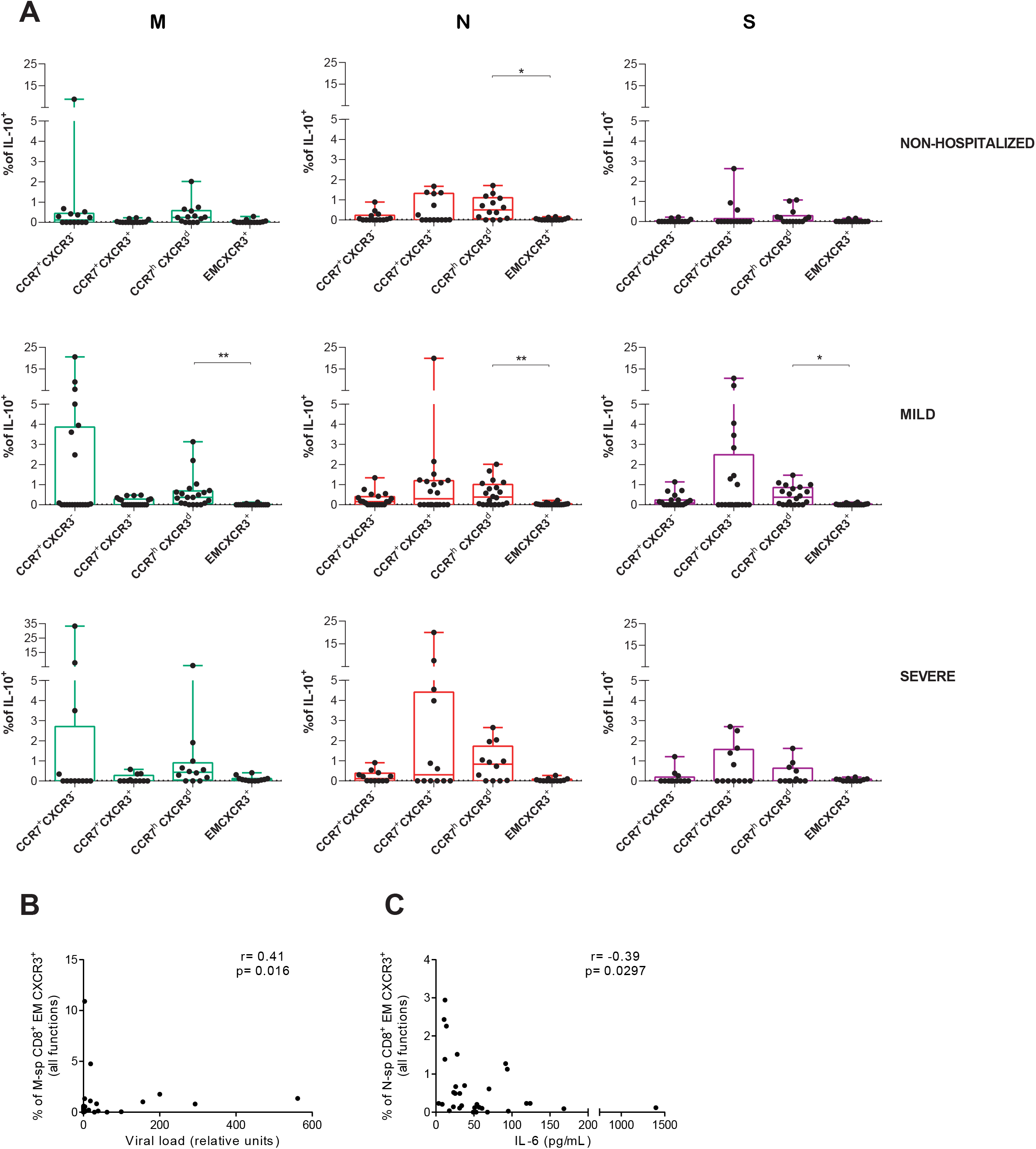
IL-10 expression in SARS-CoV-2-specific CD8^+^T cell subsets during acute infection. **(A)** Net frequency of IL-10 expression in CCR7^+^CXCR3^-^, CCR7^+^CXCR3^+^, CCR7^h^CXCR3^d^ and EM CXCR3^+^ subsets within CD8^+^ T cells after stimulation with any of the three viral proteins (membrane (M), nucleocapsid (N) and spike (S) proteins). Data are shown as median and upper range, where each dot represents an individual patient for each group (non-hospitalized n=14; mild n=20 and severe n=12). Statistical comparisons were performed using Kruskal-Wallis rank- sum test with Dunn’s multiple comparison test. *p<0.05, **p<0.01. **(B-C)** Correlation between CD8^+^ EM CXCR3^+^T cells responding with any function (added net response for IFNg, CD107a, IL-4 and IL-10) against M peptides and viral load **(B)** and against N peptides and baseline IL-6 levels **(C)**. Spearman rank correlation (n=33 for viral load and n=46 for IL-6).

### Apoptosis is enhanced in antigen-specific and non-specific T cells during severe infection

We included caspase-3 in the flow cytometry panel as a surrogate marker of apoptotic cell death activation^42^, which was quantified in both antigen-specific and bystander T cells from the different subsets of the study groups (Figure 1A). Overall expression of caspase-3 in response to stimulation was increased in total CD4^+^T cells of the severe group after M and PMA/Io stimulation (p<0.0001 and p=0.032, respectively) and in CD8^+^T cells after S stimulation in comparison to the non-hospitalized group (p=0.0009) (Figure 5A). Moreover, caspase-3 expression in CD4^+^ and CD8^+^T cells after stimulation with S-peptides positively correlated with baseline IL-6 and with the number of hospitalization days for CD8^+^T cells (Figure 5B). In addition, the overall frequency of CD4^+^T cells expressing caspase-3 in response to PMA/Io positively correlated with viral load (Figure 5B). Further, an increased expression of caspase-3 within the CCR7^h^CXCR3^d^ subset was, in general, linked to the disease severity (Figure 5C), reaching statistical significance after PMA/Io stimulation when comparing the severe and the non-hospitalized groups (p=0.031; Figure 5C). Consequently, those frequencies in response to stimulation (N, S or PMA/Io) correlated positively with the number of days at the hospital and with baseline IL-6 (Figure 5D). Expression of caspase- 3 in other CCR7^+^CD8^+^T cell subsets in response to N and S peptides also correlated with IL-6 baseline levels in patients (Figure 5D).

**Figure 5.**
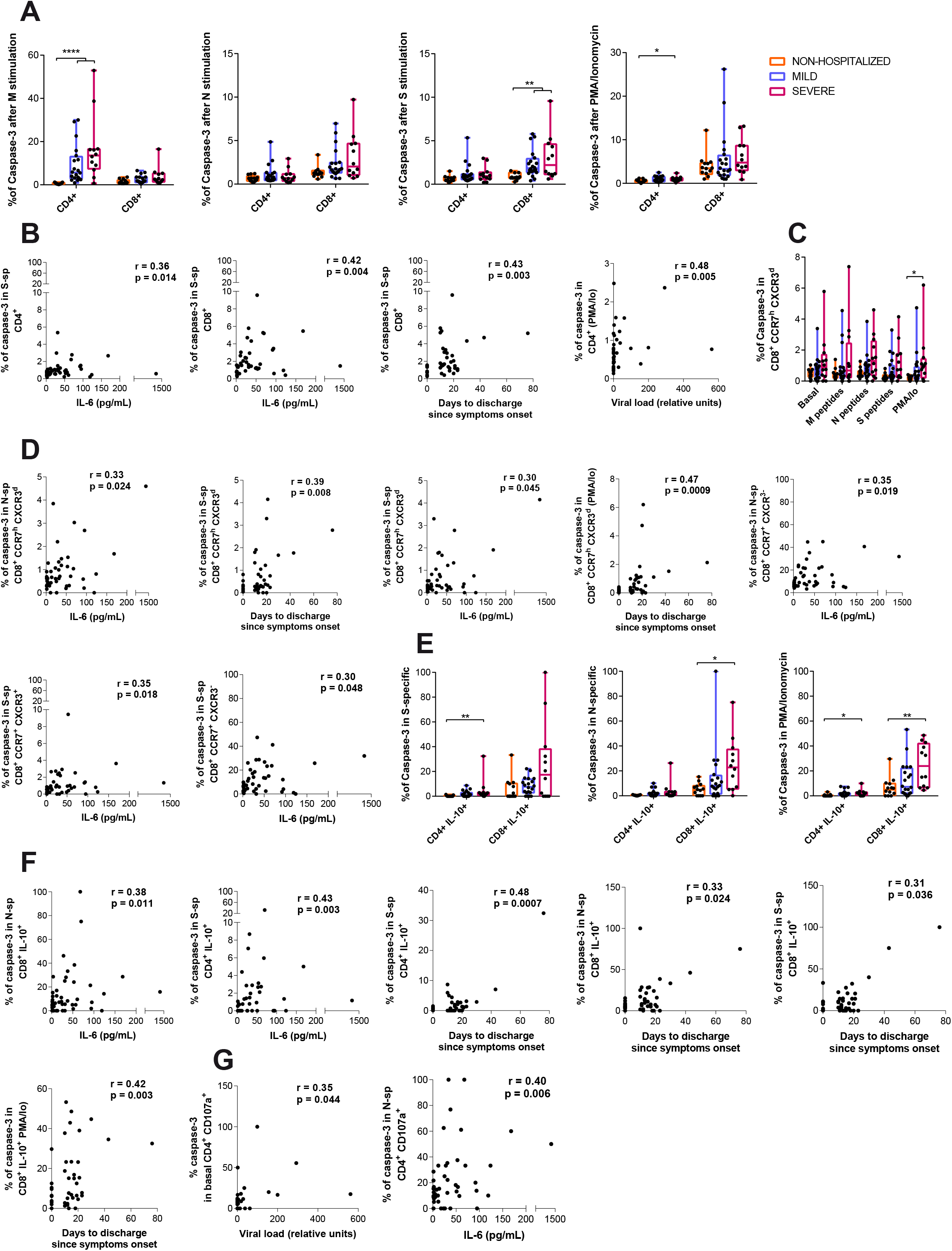
Pro-apoptotic caspase-3 expression in T cells during acute SARS-CoV-2 infection. **(A)** Frequency of caspase-3 expression in CD4^+^ and CD8^+^T cells after stimulation with membrane (M), nucleocapsid (N) or spike protein (S) and PMA/Ionomycin, in non-hospitalized (orange, n=14), mild (purple, n=20) and severe (pink, n=12) COVID-19 patients. **(B)** Correlation between days to hospital discharge since symptoms onset, viral load or baseline IL-6 levels (pg/mL) and the net frequency (background subtracted) of caspase-3 in CD4^+^ and CD8^+^T cells after stimulation with the spike protein or PMA/Ionomycin. **C)** Frequency of caspase-3 expression in CD8^+^ CCR7^h^CXCR3^d^T cells after stimulation. **(D)** Correlations between clinical parameters and the net frequency of caspase-3 expression in CD8^+^ CCR7^+^ T cell subsets after stimulation. **(E)** Frequency of caspase-3 expression in IL-10-secreting SARS-CoV-2-specific CD4^+^ and CD8^+^T cells responding to the spike protein, the nucleocapsid protein or to PMA/Ionomycin. **(F)** Correlation between clinical parameters and IL-10-expressing SARS-CoV-2-specific CD4^+^ or CD8^+^T cells, or after PMA/Ionomycin stimulation. **G)** Correlation between viral load and the frequency of caspase-3 expression in basal CD107a^+^ degranulating CD4^+^T cells and between IL-6 and the net frequency of caspase-3 expression in CD107a-expressing CD4^+^ in response to N peptides. Data in graphs are shown as median and min to max range and statistical comparisons were performed using Kruskal-Wallis rank-sum test with Dunn’s multiple comparison test. *p<0.05, **p<0.01, ***p<0.001, ****p<0.0001. Spearman rank correlation (n=46 for all correlations except for viral load which corresponds to n=33).

Regarding antigen-specific T cells we detected remarkable differences within the IL-10 secreting T cells. In this sense, increased expression of caspase-3 was distinguished in the hospitalized severe group compared to the non-hospitalized in S-specific IL-10^+^ CD4^+^T cells (p=0.004), N-specific IL-10^+^ CD8^+^T cells (p=0.016) and even in the overall IL-10 secretion capacity in response to PMA/Io response (p=0.031 for CD4^+^ and p=0.006 for CD8^+^; Figure 5E). Further, significant correlations between apoptosis in IL-10 antigen-specific T cells and several clinical parameters supported these results (Figure 5F). As for the other functions, we only detected positive correlations for the expression of caspase-3 in baseline and N-specific CD107a^+^ CD4^+^T cells with viral load and IL-6 levels, respectively (Figure 5G). These results indicate increased activation-induced cell death associated with viral replication and disease severity affecting total CD4^+^ and CD8^+^T cells, a phenomenon that seems to be modulated by the viral protein targeted. Moreover, CD8^+^CCR7^h^CXCR3^d^ T cells, a major producers of IL-10, appeared to be one of the most affected population.

### Ag-specific T_RM_ responses are present in the lung of convalescent patients

In order to demonstrate that antigen-specific T cells detected during acute SARS-CoV-2 infection, not only migrate into the lung parenchyma but also persist as T_RM_, we measured their frequency in lung biopsies of five patients. These patients, who strongly differed in their SARS-CoV-2 infection profile, successfully recovered and SARS-CoV-2 was not detected in the respiratory tract by RT-PCR before they underwent thoracic surgery for different reasons. Briefly, HL24 patient was a young-asymptomatic patient whose blood and lung samples were analyzed 21 days after SARS-CoV-2 laboratory-confirmation by RT-PCR. In contrast, samples were analyzed between 6 and 10 months after initial SARS-CoV-2 RT-PCR confirmation for the two mild cases (HL52 and HL65) and the two severe cases (HL27 and HL69). Of note, a more detailed COVID-19 clinical history can be found in the methods section.

Antigen-specific T cell responses were analyzed in total lung CD4^+^ and CD8^+^T cells and by three different fractions: CD69^-^ (non-T_RM_), CD69^+^ (T_RM_) and a subset within CD69^+^ cells expressing CD103^+^ (T_RM_ CD103^+^) (Figure S7A). Of note, CD69^+^T cells did not express T-bet confirming their T_RM_ nature^20^. As shown for one of the convalescent patients with previous severe disease (HL27; Figure 6A), antigen-specific T cells secreting IFNg were restricted to the T_RM_ fractions, which in the case of S-specific CD4^+^T cells represented up to 3.47% of the T_RM_ CD103^+^ subset. Similarly, in the lung biopsy of a mild convalescent patient (HL52; Figure 6B) N-specific CD8^+^T cells secreting IFNg were restricted to the T_RM_ fractions and >40% of them were simultaneously degranulating (CD107a^+^). In fact, the assessment of the frequency of cells responding to each peptide pool demonstrated a general trend for more antigen-specific T cells belonging to the T_RM_ subsets for all functions and all patients (Figure 6C and S7B-F). Regardless of the variability observed among study patients, the frequency of specific T cell responses increased with disease severity (from left to right; Figure 6C), while their magnitude was in general low and less consistent for IL-4 or IL-10 responses (Figure 6C and S7B-F). Importantly, a consistent polyfunctional IFNg^+^CD107a^+^ T cell response, which represented between 0.025 and 0.051% of all CD3^+^T cells and was mostly associated with the T_RM_ fraction (>75%), was detected against N peptides in all patients except in the asymptomatic one (HL24; Figure 6D).

**Figure 6.**
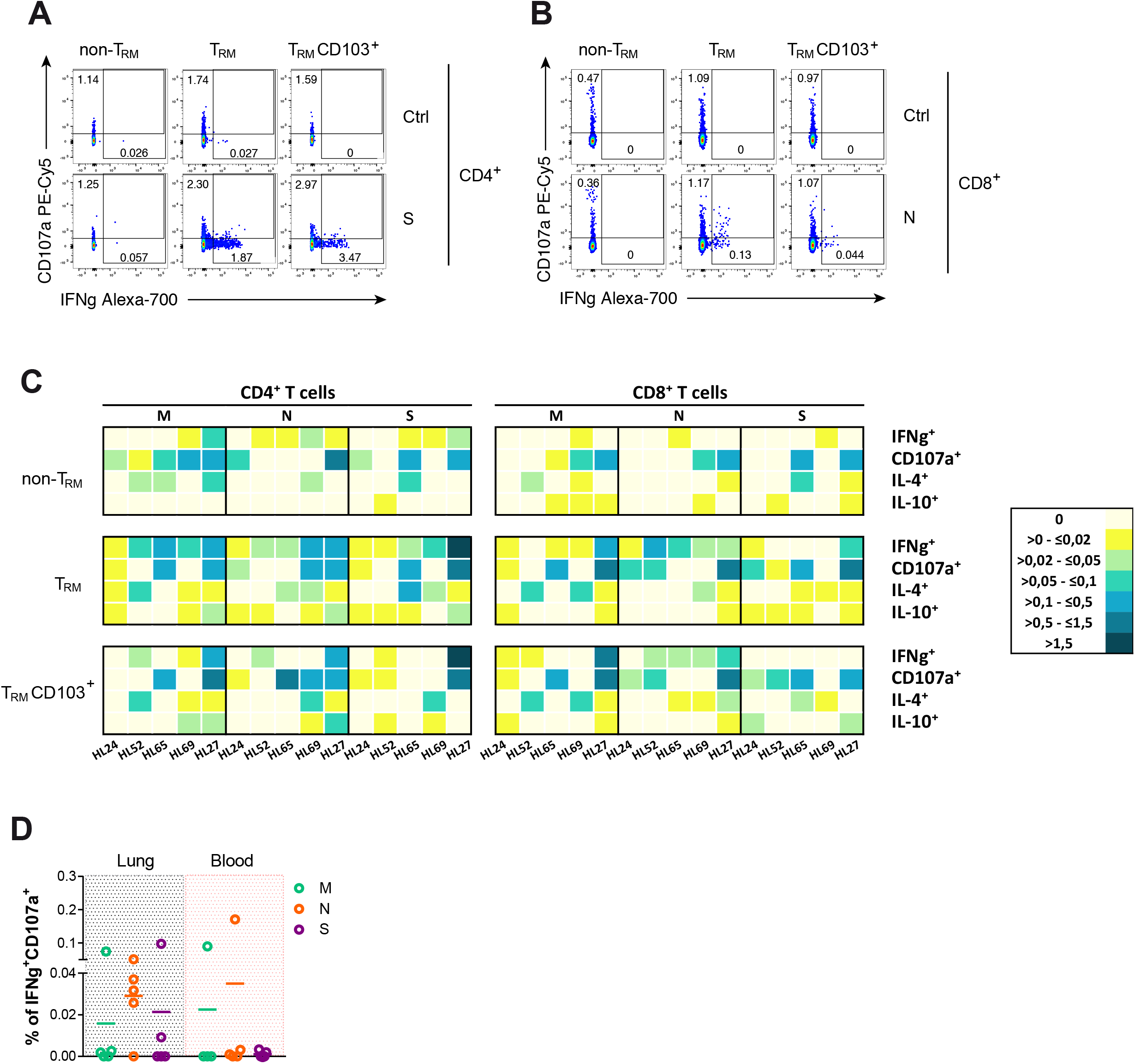
Functional analysis of lung-resident SARS-CoV-2-specific T cells. **(A and B)** Flow cytometry plots showing the frequency of IFNg and degranulation (CD107a) by non-T_RM_, T_RM_ or CD103^+^ T_RM_ in CD4^+^ from HL27 after spike stimulation and control **(A)** and in CD8^+^ from HL52 after nucleocapsid stimulation and control **(B). (C)** Heatmaps summarizing the net frequencies of IFNg, CD107a, IL-4 and IL-10 SARS-CoV-2-specific CD4^+^ or CD8^+^ non-T_RM_, T_RM_ and T_RM_ CD103^+^ from 5 different SARS-CoV-2 recovered patients. Cytokine production or degranulation are displayed as colors ranging from yellow to blue, based on the frequency, as shown in the key. **(D)** Net frequency of double positive IFNg/CD107a CD3^+^T cells from lung or blood after stimulation with membrane (M; green), nucleocapsid (N; orange) or spike protein (S; purple.

The comparison between the overall SARS-CoV-2-specific T cell responses detected in lungs with the ones found in contemporary peripheral blood samples highlighted strong differences between these two compartments (Figure 7). For example, in the asymptomatic HL24- patient IFNg and IL-10 responses were more frequent in blood than in lung, IL-4 was absent, and T cell degranulation was only detectable in lung (Figure 7A). Importantly, this patient was closer to the initial RT-PCR-based laboratory confirmation (3 weeks after), and his profile was more consistent with a non-hospitalized patient. A different pattern was observed for one of the severe convalescent patients (HL27), who had persistent T_RM_ in the lung overrepresented by S-specific CD4^+^T cells secreting IFNg, which represented up to 1.58% of the total CD4^+^T cells, while only 0.082% of the circulating CD4^+^T cells secreted IFNg and, in this case, in response to M peptides (Figure 7E). In fact, whereas all four functions were detected in T cells from lung after 6 months since initial infection for this patient, they were barely detectable in blood. Overall, viral-specific T_RM_ responses were detected in all patients. However, no consistent patterns were observed among patients in terms of viral proteins targeted and functions between blood and lung compartments, except for the polyfunctional response detected in tissue against N peptides. Of note, the asymptomatic patient had no detectable antibodies, whilst the two severe and two mild patients had detectable antibodies against SARS-CoV-2 in a concomitant plasma sample (measured as total and as IgG fraction). Furthermore, no virus was detected in any of the lung biopsies by immunofluorescence or viral RNA in situ hybridization. Overall, our results highlight the establishment and persistence of lung-resident T cell immunity against SARS-CoV-2 viral infection. However, in most cases, antigen-specific T_RM_ patterns could not be identified in T cells from blood.

**Figure 7.**
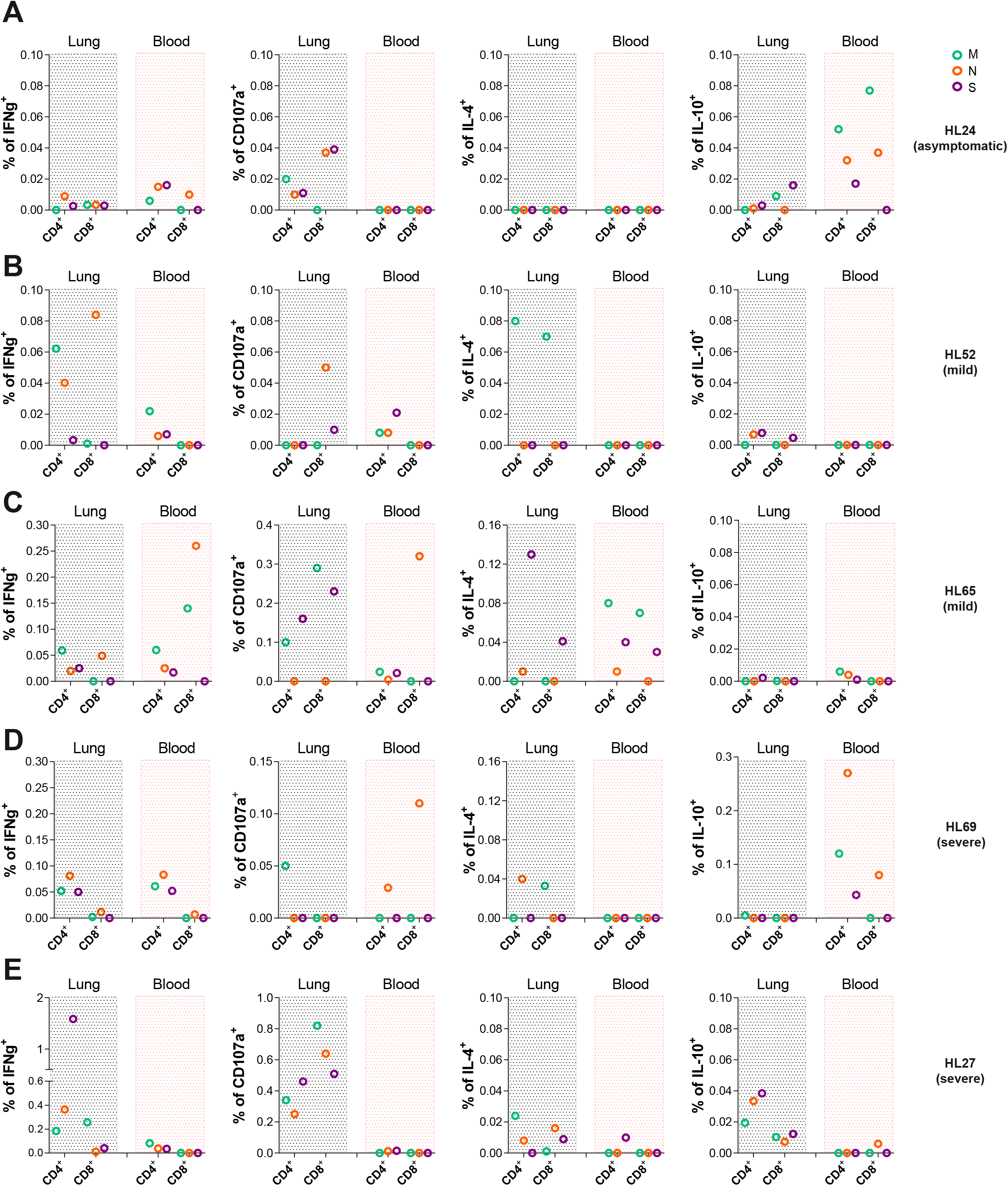
Comparison between SARS-CoV-2-specific T cells in lung and blood of convalescent patients. **(A-D)** Total CD4^+^ and CD8^+^ T cell net frequencies of IFNg, CD107a, IL-4 and IL-10 expression in SARS-CoV-2-specific T cells derived from lung or blood from the same patient **(A)** (HL24), **(B)** (HL52), **(C)** (HL65), **(D)** (HL69) and **(E)** (HL27). Viral proteins are shown in color green (membrane protein, M), orange (nucleocapsid protein, N) and purple (spike protein, S).

## DISCUSSION

This study identifies unique features of the cellular immunological response against SARS-CoV- 2 relevant to infection control and disease progression, which may be critical to informing vaccine assessment and development of new prototypes. First of all, we show that the acute response of non-hospitalized infected patients is characterized by CD4^+^ and, to less extent, CD8^+^ SARS-CoV- 2 specific T cells secreting IL-10, to which subsets expressing high levels of CCR7 contribute abundantly. In contrast, hospitalized patients show a bias towards an effector response characterized by IFNg and IL-4 secretion, being the main functions as severity increases. Second, depending on the SARS-CoV-2 viral protein targeted, different CD4^+^ and CD8^+^T cell functional profiles are generated, which have clear implications for vaccine design. Third, lymphopenia is partially a consequence of increased apoptosis in antigen-specific and non-specific T cells, which is associated with disease severity and where SARS-CoV-2 specific subsets, such as IL-10 secreting T cells, appear to be more susceptible. Lastly, and most important, SARS-CoV-2 T_RM_ can be established and persist for 10 months after infection; nonetheless, the magnitude and profile of the lung SARS-CoV-2 specific T cells strongly differ from the response detected in blood.

Major efforts have recently centered on the identification and characterization of SARS-CoV-2 specific T cells^4, 8, 9, 10, 11, 12, 13, 14^. Several of these studies have focused on defining the viral proteins more often targeted by specific T cells, concluding that after infection a broad cell response against multiple structural and non-structural regions of SARS-CoV-2 is detected in most convalescent patients^4, 9, 12^. More recently, it has been reported that SARS-CoV-2 specific T cells appear to be weaker and less frequent during acute infection^13^. In this sense, in our study, while the frequency of responders based on CD4^+^T cells specifically secreting IFNg was similar to previous reports, they were indeed weak in terms of the amount of IFNg or cytotoxicity. However, SARS-CoV-2-specific T cell response during acute infection appeared to be dominated by IL-4 secretion in hospitalized patients and by IL-10 in non-hospitalized patients. A recent report highlights that, in contrast to other cytokines, increased levels of SARS-CoV-2 specific CD4^+^T cells secreting IL-10 were mainly detected during active disease^43^. However, these functions have rarely been assessed as part of the specific intracellular T cell response, and others have measured IL-4 in the supernatant of stimulated PBMC without detecting any increase^9, 13, 14^. In contrast, higher serum levels of IL-4, IFNg and IL-10 cytokines, among others, have been associated with COVID-19^29, 36, 37, 39^. Differences in methodology and sample timing may account for these discrepancies. In fact, we did not detect changes in plasma levels of IL-4, highlighting differences between measuring the overall level of a given cytokine released systemically rather than the capacity of a small frequency of antigen-specific T cells to quickly secrete such cytokine. Importantly, several studies have detected a negative impact of IL-4 mediated responses on immune protection^44^. Furthermore, an increased expression of IL-4 was detected in the lungs of patients who died from SARS-CoV-2 infection^45^. In our cohort, spontaneous IL-4 secretion from both T cell subsets correlated with disease severity, and responses against the spike protein strongly stimulated this response, potentially suggesting the induction of a stronger antibody-directed response. Last, the patient with mild infection who tested positive for 4 months (HL65) showed the most frequent IL-4 response in the lung, which was against S peptides.

Caspase-mediated apoptosis in the immune system is a major contributor to immune homeostasis in a process termed activation-induced cell death^46^, which could potentially contribute to minimize an overwhelming cytokine response. Our results are consistent with apoptosis significantly contributing to the lymphopenia detected in COVID-19 patients, and the preferential loss of CD8^+^T cells is accompanied by an increase in caspase-3 within this compartment, consistent with a pre-publication^47^. Since inflammatory molecules can often be potent activators of cell death, increasing levels of IL-10 may moderate the extent of apoptosis induced, as occurs in mouse models of bacterial infection^48^. Moreover, not only may the inflammatory environment contribute to a higher proportion of bystander T cells succumbing to apoptotic cell death, but viral proteins may also induce apoptosis in both antigen and non-antigen specific T cells. While effector T cells with degranulation capacity are expected to be more terminally differentiated and, consequently, may be more prone to activation-induced cell death, the fact that other subsets (i.e. CCR7^h^CXCR3^d^/IL-10 secreting) were more affected is intriguing and requires further study.

Critically, the only cytokine that was higher in non-hospitalized compared to hospitalized groups was IL-12p70 (Figure S1B), suggesting that, as occurs with other respiratory viruses^49^, cell-mediated Th1 immunity is necessary for recovery from respiratory infection. A general predominance of a Th1 profile does not compromise the generation of neutralizing antibodies^44^. Further, it is increasingly accepted that Th-cell subsets are plastic, especially during responses to pathogens *in vivo*, and even cytokines such as IL-10 can be produced by subpopulations of cells within multiple effector subsets^44^. However, SARS-CoV-2-specific T cells from acute responders demonstrated a biased Th1 profile, where IFNg-secreting T cells could also secrete IL-2 in the case of CD4^+^ and TNFa and granzyme B, but not IL-10, in the case of CD8^+13^. Our data also shows marginal co-expression between IFNg, IL4 and IL-10, suggesting multiple polarizations of antigen-responding T cells^44^, which is also supported by different CCR7/CXCR3 subsets being the main contributors to a given response. Moreover, this polarization was partially induced by the targeted protein, where M peptides induced the strongest IFNg secretion in CD4^+^T cells, N peptides enhanced cytotoxicity in CD8^+^T cells and S peptides had an overall predominant Th2 profile (IL-4). Similar to what was observed for convalescent patients^9^, M and N-specific responses dominated in non-hospitalized and mild hospitalized cases, indicating that a candidate vaccine including only SARS-CoV-2 spike would limit the array of responses during natural infection^9^. Furthermore, a comparison between the functional profile of antigen-specific CD4^+^ and CD8^+^T cells based on the viral protein targeted evidenced a wider functional profile for CD8^+^T cells targeting M or N peptides compared to S, which was more often observed in milder cases than in severe ones^12^ and agrees with recent identification of responses to frequently recognized CD8^+^T cell epitopes^50^. Overall, our results concur with a broader Th profile in CD8^+^T cells induced by the non-spike viral proteins and associated with a less severe infection, while spike responses dominated by IL-4^+^ CD8^+^T cells represented a hallmark of disease severity, being the sole response in the fatal case. Further, the observation that N-specific EM CXCR3^+^CD8^+^T cell responses with an IFNg^+^CD107a^+^ Th1 profile were associated with non-hospitalization during symptomatic COVID-19 suggests a favorable immune response when the N protein is targeted by CD8^+^T cells and these responses can migrate towards infected tissues. The CXCR3-CXCL10 axis appears critical for the recruitment of CD4^+^ and CD8^+^T cells that control influenza and tuberculosis infection in the lung respectively^25, 51^. Consequently, lung-T cell recruitment may partially contribute to the lymphopenia detected in patients^52^, where this antiviral response will likely establish as resident memory cells. In fact, N-specific IFNg^+^CD107^+^ T_RM_ were detected in all four convalescent patients 6 to 10 months after infection.

Recent deep immune profiles of COVID19 patients have identified T-bet expression as a transcriptional factor associated to patients with better prognosis^53^. Importantly, T-bet is not only a key regulator of Th1 immune effector responses and CXCR3 expression, essential for effective clearance of pathogens and maintenance of immunity^54^, but also crucial for migration, proliferation and survival of T regulatory cells (Treg) during Th1-mediated immune responses *in vivo*^55^. Indeed, CXCR3 is also found on a subset of CD4^+^Foxp3^+^T cells, and the control of inflammatory responses at mucosal surfaces requires IL-10 producing Treg^55^. One of the main correlates of disease control during acute infection was IL-10 secretion, which dominated the specific immune response of non-hospitalized patients. Two previous studies have detected reduced frequencies of Treg in severe COVID-19 cases^26, 56^; however, IL-10 was detected in supernatants from stimulated PBMC of severe patients during acute infection^14^, and a similar trend was observed in a cohort of acute patients with a wider range of sampling days, 4-37^13^. While, as detected in the plasma of our own cohort of patients, increased serum levels of IL-10 have been widely associated with COVID-19 and disease severity (reviewed in^37^), several factors may explain these results. The plasma source of IL-10 can have multiple cellular origins, ranging from myeloid subsets to epithelial cells, since not only may almost all leukocytes produce IL-10, but also the range amplifies during inflammation^48^. The systemic increase in IL-10 may still act as a compensatory response to limit massive ongoing inflammation in severe patients^57^. We propose that an early effector specific T cell response coordinated with engagement of other immune profiles limiting inflammation may aid at promoting infection resolution. This notion is supported by comprehensive analyses of common immune correlates of protection from mortality in mouse models of influenza and SARS-CoV infection, which revealed a unique T regulatory suppressive profile that contributed to this balance^58^. Furthermore, it has previously been shown that Treg activity is required during viral infections to allow for appropriate generation and migration of immune effector cells to the site of infection^59, 60^, while blocking the action of the IL-10 secreted by antiviral T cells results in enhanced pulmonary inflammation and lethal injury^17, 48, 61^. In fact, during influenza infections, type I IFN signaling may contribute to IL-10-producing lymphocyte recruitment to the site of infection to moderate excessive inflammation, which will be coincident with the onset of the adaptive immune response^48^, being CD8^+^T cells a primary source of IL-10 production in the respiratory tract^62^. Accordingly, while IL-10 plasma levels increased with disease severity, the fatal case had one of the lowest levels in plasma, which was accompanied by an absolute lack of IL-10 secretion by antigen-specific T cells.

T_RM_ strategically residing in peripheral tissues are key to controlling mucosal infections and providing rapid and durable immunity against reinfection^20, 63^. Previous studies in SARS- recovered patients already pointed towards persistence of a memory T cell response for up to 6 years after infection, and suggested vaccine-mediated induction of T_RM_ as a long-term protection strategy^5^. In concordance, a larger proportion of CD8^+^T cell effectors with T_RM_ characteristics were present in bronchoalveolar lavages from patients with moderate infection compared to severe- infected patients^64^. We indeed report the existence of a high frequency of T_RM_ in the lung of a patient who was infected almost 6 months before, yet had a severe and durable infection (HL27). In this patient, while all functions were represented in T_RM_ in a remarkably higher proportion than in blood, IFNg in response to S peptides dominated. In this sense, particularly high frequency of spike protein-specific CD4^+^T cell responses was observed in blood in patients who had recovered from COVID-19^4, 9, 12^. Importantly, CD4^+^T cells are necessary for the formation of protective CD8^+^T_RM_ during influenza infection, and cytokines, such as IFNg, are necessary signals for this process^20^. However, CD4^+^T cells themselves can be cytotoxic and, actually, have been shown to confer protection against influenza^20^. We also detected degranulation in response to viral peptides in CD4^+^ and even more so in CD8^+^T cells from the lung, which in the case of the asymptomatic young patient were completely absent in blood. Lack of degranulation in blood from convalescent patients has also been reported^12^. Further, the fact that the lung biopsy of this young asymptomatic patient was at 3 weeks RT-PCR laboratory-confirmation of infection, suggests early recruitment of cytotoxic T cells to the lung even in asymptomatic cases. Moreover, the most frequent responses by circulating T cells from this asymptomatic patient were CD4^+^ and CD8^+^T cells secreting IL-10, which concurs with the dominating pattern in non-hospitalized patients during acute infection.

We acknowledge that our study has several limitations, one being that sample size for the different groups was small to be conclusive. However, this was compensated by a narrow window of sampling during acute infection (7-16 days, post-symptoms onset) and by a comprehensive clinical characterization to stratify patients to study groups. In this sense, multiple correlations support our main findings and provide strength to our data, which is also largely supported by current literature. Further, identification of the precise phenotypes generating antigen-specific T responses, such as Tregs for IL-10, or the consideration of other T lymphocytes such as gdT cells should also be considered in future studies. Last, only five lung biopsies obtained from very different COVID-19 convalescent patients could be studied. While these patients are so far scarce, the immune responses identified in those samples not only contributed to round out the present report, but also represent the first evidence to our knowledge of persisting SARS-CoV-2- specific T_RM_ in the lung. Disease severity during acute SARS-CoV-2 infection is associated with strong peripheral T and B cell responses^4, 27^, which not only may relate to antigenic burden but, also could potentially translate into a significant proportion of antigen-specific T_RM_ in the lung once the patient recovers. Remaining important questions concern the level of viral replication and associated symptomatology that will stimulate an effective immune response at the respiratory tract, and also, how quick this response will be established. However, the fact that an asymptomatic patient had degranulating antigen-specific T cells in the lung 3 weeks after infection is at least encouraging. Overall, a balanced effector/anti-inflammatory response may be key for early viral containment, where antigen-specific IL-10^+^T cells could be determinant in limiting inflammation. Thus, the possibility that overstimulated pro-inflammatory T cells contribute to disease severity cannot be ruled out. Our findings encourage next-generation vaccine designs to consider including viral proteins beyond the spike protein, in particular nucleocapsid peptides, which should broaden and balance the functional profile of memory T cells, resembling control of natural infection.

## METHODS

### Ethics statement

This study was performed in accordance with the Declaration of Helsinki and approved by the corresponding Institutional Review Board (PR(AG)192/2020 and PR(AG)212/2020) of the Vall d’Hebron University Hospital (HUVH), Barcelona, Spain. Written informed consent was provided by all patients recruited to this study and samples were prospectively collected and cryopreserved in the Vall d’Hebron Research Institute.

### Healthy donors

Blood samples from healthy adult donors were obtained via phlebotomy. These blood samples were collected for studies unrelated to COVID-19 between September 2018 and June 2019 (PR(AG)116/2018 and PR(AG)117-2018). At the time of enrollment in the initial studies, all individual donors provided informed consent that their samples could be used for future studies. These samples were considered to be from unexposed controls given that SARS-CoV-2 emerged as a novel pathogen in December 2019 and these samples were largely collected before this date. These donors were considered healthy in that they had no known history of any significant systemic illnesses. The cohort of healthy donors includes 12 individuals.

### Patients with SARS-CoV-2 infection

Adult patients, 18 years old and older, diagnosed with acute COVID-19 were recruited at the Vall d’Hebron Hospital during the first COVID-19 outbreak between March and May 2020. Diagnosis of acute COVID-19 was defined by symptomatology and/or clinical findings and confirmed by positive reverse-transcriptase polymerase chain reaction (RT-PCR) for SARS-CoV-2 in a respiratory tract specimen. Immunocompromised patients in which the immune response may be affected were excluded of the study. 20 milliliters of blood were collected at baseline by phlebotomy in two EDTA tubes and stored at room temperature briefly prior to processing for PBMC and plasma isolation. Samples were obtained between 7 and 16 days after symptoms onset.

Biochemistry analyses were measured at baseline in all patients and, subsequently, according to the clinical care needs of each patient during infection. Routine clinical laboratory analyses included complete blood count, coagulation testing (including D-dimer measurement), liver and renal function, electrolytes and inflammatory profile (including C-reactive protein, fibrinogen, ferritin and IL-6). The study cohort consisted of 12 patients with severe disease, 20 with mild disease and 14 non-hospitalized individuals. Patient information is summarized in Table S1. According to disease severity patients, at the discretion of the treating physician, patients were classified in three groups:

a. Patients with severe disease: individuals with radiologically confirmed pneumonia that required hospitalization and had acute respiratory failure and/or analytical parameters of severity and/or extensive radiological involvement.
b. Patients with mild disease: individuals with radiologically confirmed pneumonia that required hospital admission but without criteria of severity.
c. Non-hospitalized patients: individuals without pneumonia and with pauci-asymptomatic disease that did not require hospitalization and managed on an outpatient clinic.

Data were collected prospectively from the medical charts of the patients. We collected sociodemographic characteristics, past medical records, Charlson comorbidity score, concomitant medication, treatments against SARS-CoV-2 infection, adverse drug events, blood test results, imaging studies, microbiological tests and supportive measures needed. Vital signs, symptoms and physical examination were recorded. Laboratory, microbiology and imaging studies were performed according to the clinical care needs of each patient.

### Lung biopsies

Lung biopsies were obtained from five patients recovered from SARS-CoV-2 infection who needed a lung resection. Analyses were performed using healthy areas from the lung resection. HL24 sample corresponded to a young smoking man (in his early 20’) who had detectable viral load by RT-PCR without symptoms, followed by two negative RT-PCR measurements (8 and 18 days after the positive result). He underwent surgery for pneumothorax 21 days after the positive SARS-CoV-2 detection. HL52 corresponded to an ex-smoker man in his late 70’ who was hospitalized for five days due to symptomatology compatible with SARS-CoV-2 infection (with confirmatory RT-PCR 3 days after hospitalization and a baseline IL-6 of 26.15 pg/mL). During a post-COVID examination, a lung carcinoma was diagnosed, which instigated thoracic surgery 7.5 months after hospital discharge. HL65 was a woman in her early fifties hospitalized for three days (without oxygen requirements), with confirmatory SARS-CoV-2 RT-PCR and a baseline IL-6 of 5.25 pg/mL. During the next 4 months, she tested positive for RT-PCR. She underwent thoracic surgery for a pulmonary nodule 2 months after testing negative for RT-PCR (and 6 months after initial discharge). HL69 was a man in his late sixties hospitalized for severe COVID-19 for 35 days (baseline IL-6 of 34.66 pg/mL) and treated for the symptomatology derived of the SARS-CoV-2 infection (with confirmatory RT-PCR). During a post-COVID examination, a pulmonary nodule was diagnosed, which instigated thoracic surgery 10 months after initial infection. HL27 was an ex-smoker man in his late sixties who was hospitalized for one month due to respiratory insufficiency caused by SARS-CoV-2 infection (with confirmatory RT-PCR and a baseline IL-6 of 133.7pg/mL). One month after discharge, he presented a rash in photoexposed skin, potentially related to persistent SARS-CoV-2 infection^65^ or treatment with hydroxychloroquine<u>^66^</u>. RT-PCR for SARS-CoV-2 tested positive again, turning into a RT-PCR negative two weeks after. During this time, a lung carcinoma was diagnosed, which instigated thoracic surgery 3 months after the negative RT-PCR (and 5 months after initial discharge).

Concomitant to the lung biopsy, blood samples were also collected. Thus analyzed paired blood and tissue samples corresponded to 21 days after the first positive SARS-CoV-2 detection for the HL24 patient (asymptomatic) and between 6 and 10 months after initial infection for HL65 (mild), HL52 (mild-PCR+ for 4 months), HL69 (severe) and HL27 (severe-PCR+ for 2 months) patients. Blood samples were immediately processed for PBMC and serum isolation, which was used for SARS-CoV-2 antibody detection.

### SARS-CoV-2 RT-qPCR

Upper (naso/oropharyngeal swabs) and lower (bronchoalveolar lavage, tracheal aspirate, sputum or bronchoaspirate) respiratory tract specimens from subjects with suspicion of COVID-19 were received and tested at the Respiratory Viruses Unit of the Microbiology Department of the HUVH. COVID-19 diagnosis was performed by two commercial RT-PCR-based assays, Allplex™ 2019- nCoV (Seegene, Korea) or Cobas® SARS-CoV-2 (Roche Diagnostics, USA) tests. In addition, an in-house PCR assay using the primer/probe set targeting the nucleocapsid protein (N1) and the human RNase P (housekeeping gene), from the CDC 2019-nCoV Real-Time RT-PCR Diagnostic Panel (Qiagen, Hilden, Germany), was performed. In order to minimize variations due to a non-standardized collection of a heterogeneous specimen, the Ct values of the viral target were normalized to the housekeeping gene based on the 2^−ΔCt^ method, where ΔCt corresponds to the formula Ct sample – Ct housekeeping.

### Plasma cytokine determinations

Plasma obtained from our three cohorts of COVID-19 patients (n=45) was analyzed using Ella^®^ platform (Bio-Techne, Minneapolis, Minnesota, USA) for the quantification of the following cytokines and chemokines: CCL2, GM-CSF, IL-10, IL-12 p70, IL-1ra, IL-6, IL-7, TNFa, CXCL10, Granzyme B, IFNg, IL-13, IL-15, IL-17A and IL-4. Samples were 1:2 diluted with sample diluent provided by the manufacturer and loaded onto multiplex cartridges according to manufacturer’s instructions prior to their analysis. Results are expressed as pg/mL.

### Phenotyping and Intracellular Cytokine Staining in blood

PBMCs were isolated from blood by density-gradient centrifugation using Ficoll-Paque and immediately cryopreserved and stored in liquid nitrogen until use in the assays. Cells were thawed the day before the assay and cultured in a T-25 flask at 37°C with RPMI 1640 (Gibco) supplemented with 10% Fetal Bovine Serum (FBS) (Gibco), 100µg/ml streptomycin (Fisher Scientific) and 100 U/ml penicillin (Fisher Scientific) (R10). Next day, previous to SARS-CoV-2 peptide pool stimulation, cells were stained for CCR7 (PE-CF594, BD Biosciences) and CXCR3 (BV650, BD Biosciences) for 30 min at 37°C. After a washing with PBS, PBMCs were stimulated in a round bottom 96-well plate for 5h at 37°C with 1µg/ml of SARS-CoV-2 peptides (PepTivator SARS-CoV-2 M, N and S, Miltenyi Biotec) in the presence of 1 μl/ml of Brefeldin A (BD Biosciences), 0.7 μl/ml of Monensin (BD Biosciences) and 3 μl/ml of α-CD28/CD49d (clones L293 and L25, BD Biosciences). Anti-CD107a (PE-Cy7, BD) was also added at this time. For each patient, a negative control, cells treated with medium, and positive control, cells incubated in the presence of 81nM PMA and 1μM Ionomycin, were included. After stimulation, cells were washed twice with PBS and stained with Aqua LIVE/DEAD fixable dead cell stain kit (Invitrogen). Cell surface antibody staining included anti-CD3 (Per-CP), anti-CD4 (BV605) and anti-CD56 (FITC) (all from BD Biosciences). Cells were subsequently fixed and permeabilized using the Cytofix/Cytoperm kit (BD Biosciences) and stained with anti-Caspase-3 (AF647, BD Biosciences), anti-Bcl-2 (BV421, Biolegend), anti-IL-4 (PE-Cy7, eBioscience), anti-IL-10 (PE, BD Biosciences) and anti-IFNg (AF700, Invitrogen) for 30 mins. Cells were then fixed with PBS 2% PFA and acquired in a BD LSR Fortessa flow cytometer (Cytomics Platform, High Technology Unit, Vall d’Hebron Institut de Recerca). FMO controls were used to draw the gates for each function.

For the patients with lung biopsies, their contemporary blood sample was processed immediately and the T cell response assay was performed without a previous cryopreservation step. Isolated PBMC were rested for 4h in the incubator and then stimulated with the same SARS-CoV-2 peptides (M, N and S) overnight, following the same protocol described above with half the amount of Brefeldin A and Monensin to avoid toxicity.

### Phenotyping and Intracellular Cytokine Staining in lung

Lung biopsies were collected in antibiotic-containing RPMI 1640 medium from the Thoracic Surgery Service at the Vall d’Hebron University Hospital. Immediately following surgery, the tissue was dissected into approximately 8-mm^3^ blocks. These blocks were first enzymatically digested with 5 mg/ml collagenase IV (Gibco) and 100µg/ml of DNase I (Roche) for 30 min at 37 °C and 400 rpm and, then, mechanically digested with a pestle. The resulting cellular suspension was filtered through a 70µm pore size cell strainer (Labclinics), washed twice with PBS and cultivated with R10 in a round-bottom 96-well plate overnight at 37°C with 1µg/ml of SARS-CoV-2 peptides (M, N and S) in the presence of 3μL/mL α-CD28/CD49d (clones L293 and L25, BD Biosciences), 0.5μL/mL Brefeldin A (BD Biosciences), 0.35μL/mL Monensin (BD Biosciences) and 5 μL/mL anti- CD107a-PE-Cy5. For each patient, a negative control, cells treated with medium, and positive control, cells incubated in the presence of 40.5nM PMA and 0.5μM Ionomycin, were included. Next day, cellular suspensions were stained with Live/Dead Aqua (Invitrogen) and anti-CD103 (FITC, Biolegend), anti-CD69 (PE-CF594, BD Biosciences), anti-CD40 (APC-Cy7, Biolegend), anti-CD8 (APC, BD Biosciences), anti-CD3 (BV650, BD Biosciences) and anti-CD45 (BV605, BD Biosciences) antibodies. Cells were subsequently fixed and permeabilized using the FoxP3 Fix/Perm kit (BD Biosciences) and stained with anti-IL-4 (PE-Cy7, eBioscience), anti-IL-10 (PE, BD Biosciences), anti-T-bet (BV421, Biolegend) and anti-IFNg (AF700, Invitrogen) antibodies. After fixation with PBS 2% PFA, cells were acquired in a BD LSR Fortessa flow cytometer.

### SARS-CoV-2 serology

Serological status of HL24, HL27, HL52, HL65 and HL69 patients was determined in serum using two commercial chemiluminescence immunoassays (CLIA) targeting specific SARS-CoV-2 antibodies: 1) Elecsys® Anti-SARS-CoV-2 (Roche Diagnostics, USA) was performed on the Cobas 8800 system (Roche Diagnostics, USA) for qualitative determination of total antibodies (including IgG, IgM and IgA) against nucleocapsid SARS-CoV-2 protein; and 2) Liaison SARS-CoV-2 S1/S2 IgG (DiaSorin, Italy) was performed on the LIAISON® XL Analyzer (DiaSorin, Italy) for quantitative determination of IgG against the spike (S) glycoprotein subunits 1 and 2 (S1/S2).

### SARS-CoV-2 detection by Immunofluorescence and RNA hybridization

Paraffin-embedded lung tissue samples were processed and analyzed at the Pathology Department of the HUVH. For SARS-CoV-2, lung tissue sections of 3μm were deparaffinized with xylene and dehydrated in ethanol. Samples were pretreated with CC2 (pH=6), and rinsed with working PBS. SARS-CoV-2 (SARS-CoV Nucleoprotein / NP Antibody, Rabbit PAb, 6F10, Sino Biological, dilution at 1:1000) antibody was applied and incubated during 1h. After washing in PBS, slides were mounted in 80% glycerol and sealed. Images were taken using BenchMArk Ultra Ventana System.

RNA hybridization was performed using RNAscope VS Universal Assays and the Ventana Discovery Ultra System. A high sensitivity target-specific probes to SARS-CoV-2 mRNA sequence (probe V-nCoV2019-S, ADC Biotecnec biology) were used. Lung tissue sections of 3μm tissue sections were mounted on Superfrost Plus microscope slides (Fisher Scientific). The assay was performed according to manufacturer’s instructions. Briefly, samples were deparaffinized and pretreated as mentioned. Next, probes were incubated for 2h at 40°C and samples were stored overnight in 5x saline sodium citrate buffer. The following day, amplification and signal development was performed by sequential incubation of Pre-Amplifiers, Amplifiers and label probe according to the manufacturer’s instructions (Kit DISCOVERY mRNA, Roche). Lastly, samples were revealed with DAB staining (3,3′-Diaminobenzidine). The experiment controls used were infected and non-infected HeLA cells.

### Statistical analyses

Flow cytometry data was analyzed using FlowJo v10.7.1 software (TreeStar). Data and statistical analyses were performed using Prism 7.0 (GraphPad Software, La Jolla, CA, USA), unless otherwise stated. The statistical specifics of the experiments are provided in the respective figure legends. Data plotted in linear scale were expressed as median + Interquartile (IQR) or Min to Max range, unless otherwise stated. Correlation analyses were performed using Spearman rank correlation. Mann-Whitney and Wilcoxon tests were applied for unpaired or paired comparisons, respectively, while Kruskal-Wallis rank-sum test with Dunn’s post hoc test was used for multiple comparisons. A *P value* < 0.05 was considered significant. For most analyses, antigen-specific T cell data has been calculated as the net frequency, where the individual percentage of expression for a given molecule in the control condition (vehicle) has been subtracted from the corresponding SARS-CoV-2-peptide stimulated conditions.

## Data Availability

All data is included as figures and supplementary data.

## ACKNOWLEDGMENTS

We would like to thank all the patients who participated in the study. We also thank Prof. Shawn C. Kefauver for thoughtful review of the manuscript. This work was primarily supported by a grant from the Health department of the Government of Catalonia (DGRIS 1_5). This work was additionally supported in part by the Spanish Health Institute Carlos III (ISCIII, PI17/01470 and ISCIII COV20/00416), the Spanish Secretariat of Science and Innovation and FEDER funds (grant RTI2018-101082-B-I00 [MINECO/FEDER]), the Spanish AIDS network Red Temática Cooperativa de Investigación en SIDA (RD16/0025/0007), the European Regional Development Fund (ERDF), the Fundació La Marató TV3 (grants 201805-10FMTV3 and 201814-10FMTV3) and the Gilead fellowships GLD19/00084 and GLD18/00008. M.J.B is supported by the Miguel Servet program funded by the Spanish Health Institute Carlos III (CP17/00179). N.M. is supported by a Ph.D. fellowship from the Vall d’Hebron Institut de Recerca (VHIR) and A.A-G and N.S-G are supported by a Ph.D. fellowship from the Spanish Secretariat of Science and Innovation (BES- 2016-076382, PRE2019-087393). The funders had no role in study design, data collection and analysis, the decision to publish, or preparation of the manuscript.

## AUTHOR CONTRIBUTIONS

Conceptualization, MJ.B. and M.G.; Patient Recruitment and Sample Collection, J.R., A.T., B.P., J.N., P. S., AL.A., B.A., V.F., J.B.; Methodology, A.F., C.K., J.G-E., N. S-G., N.M., M. S., J.E., AN.A., and S.RC.; Investigation, J.G-E., N. S-G., N.M., M. S., A.A-G., D.P., D.A-S, I.S., J.E., AN.A. and M.G.; Formal Analysis, J.G-E., N. S-G., N.M., M. S., D. P. and M.G.; Writing-Original Draft J.G-E., N. S-G., N.M., M. S. and M.G; Writing- Review & Editing, R.P-B., MJ.B. and M.G; Funding Acquisition, MJ.B. and M.G.; all authors revised the manuscript; Supervision, MJ.B. and M.G.

