## Supplemental Figures S1 to S6 and Tables S1 and S2 for "Peripheral and lung resident T cell responses against SARS-CoV-2"

### SUPPLEMENTAL INFORMATION

#### Figure S1

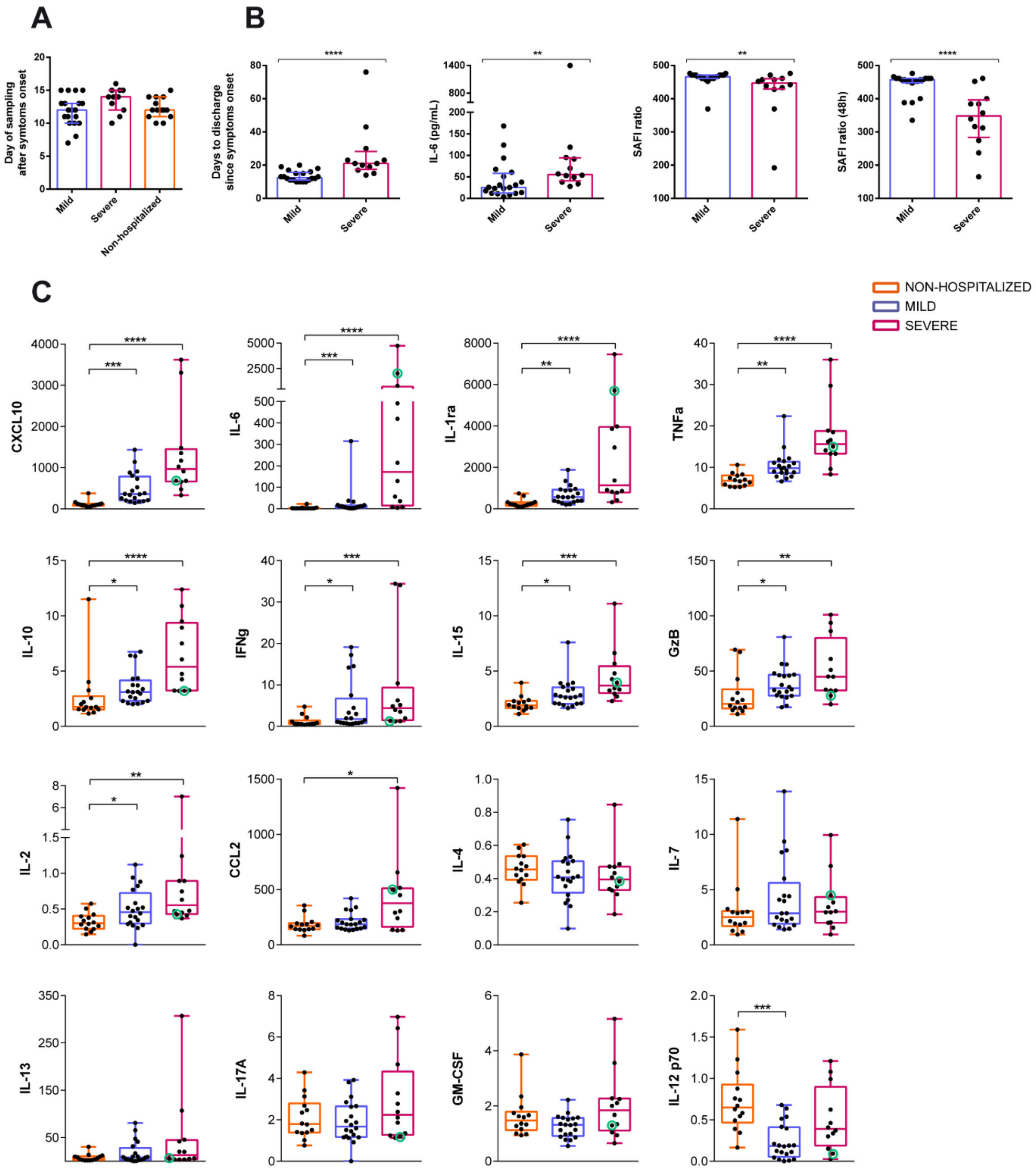

**Figure S1. Clinical parameters and plasma cytokine levels during acute SARS-CoV-2 infection.**

**(A)** Day of sample analyses by study groups (non-hospitalized n=14 in orange; mild n=20 in purple and severe n=12 in pink). Boxes and error bars represent median and interquartile range (IQR). **(B)** Pairwise comparison between mild and severe hospitalized patients for several clinical parameters: days to hospital discharge since symptoms onset, baseline IL-6 levels (pg/mL) and SAFI* ratio at baseline and after 48h. *SAFI ratio corresponds to the percentage of oxyhemoglobin saturation ($SaO_2$) in relation to the percentage of inspired oxygen ($FiO_2$). Statistical comparisons were performed using Mann-Whitney *U* tests. **(B)** Plasma levels of CXCL10, IL-6, IL-1ra, TNFa, IL-10, IFNg, IL-15, GzB, IL-2, CCL2, IL-4, IL-7, IL-13, IL-17A, GM-CSF and IL12p70 in non-hospitalized (n=14) mild (n=20) and severe (n=12) COVID-19 patients. Green circle in the severe group indicates the deceased patient. Data are shown as individual patients and median with min to max range. Statistical comparisons were performed using Kruskal-Wallis rank-sum test with Dunn's multiple comparison test. $*p < 0.05$, $**p < 0.01$, $***p < 0.001$, $****p < 0.0001$.

### Figure S2

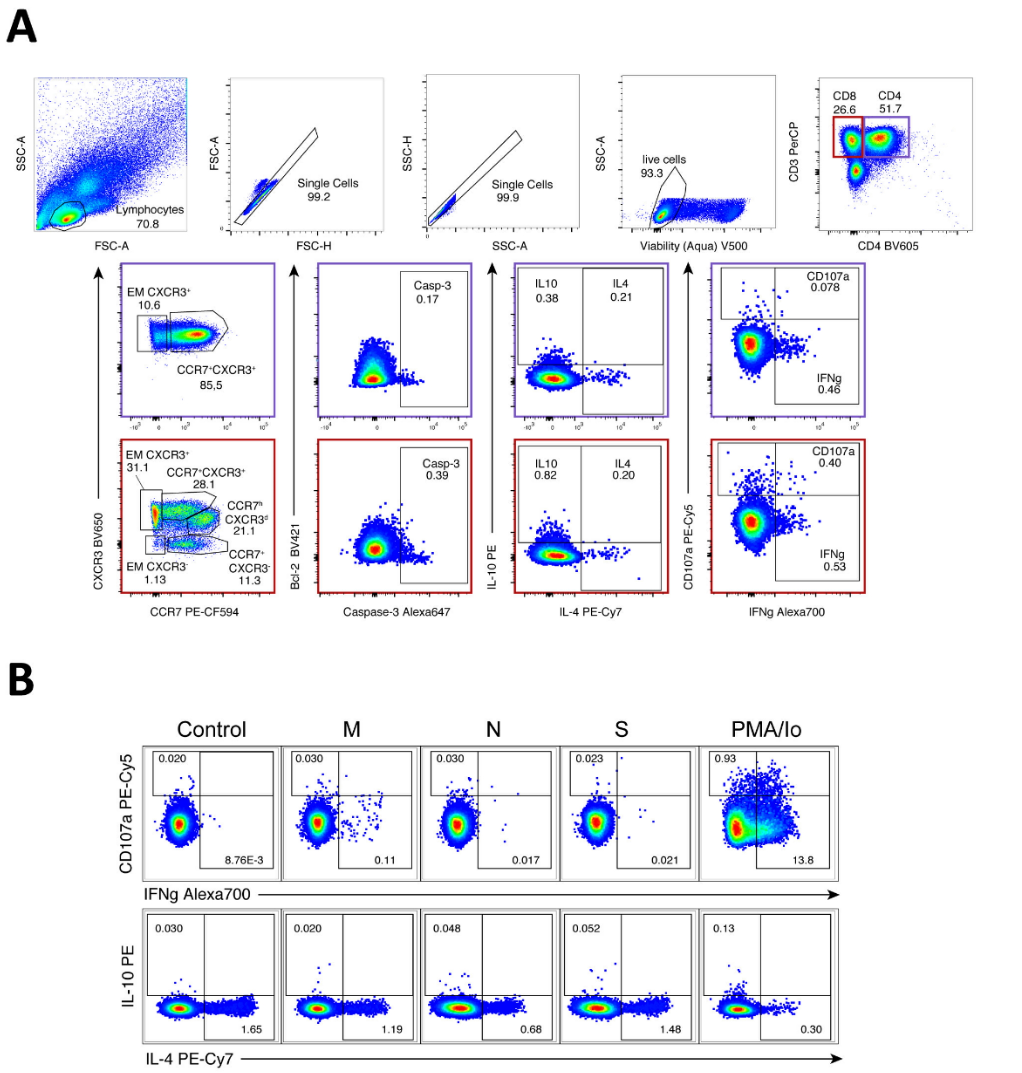


**Figure S2. Gating strategy for the analyses of SARS-CoV-2-specific $CD4^+$ and $CD8^+$ T cells. (A)** General gating strategy for functional analysis of SARS-CoV-2-specific $CD4^+$ and $CD8^+$T cell populations. Gating strategy consisted of a lymphocyte gate based on FSC vs. SSC, followed by doublet and dead exclusion, live $CD3^+$ T cell gate from where $CD4^+$ (purple) and $CD8^+$T cells were gated. Out of these two T cell subsets (below), representative subsets based on their expression of CCR7 and CXCR3, the expression of caspase-3 and the expression of diverse functions among $CD4^+$ (top) and $CD8^+$ T cells (bottom) are shown. **(B)** Representative flow cytometry plots showing individual functions after stimulation with the negative control, viral proteins (membrane (M), nucleocapsid (N) and spike (S) proteins) and PMA/Ionomycin are shown for $CD4^+$T cells from a severe patient.

### Figure S3

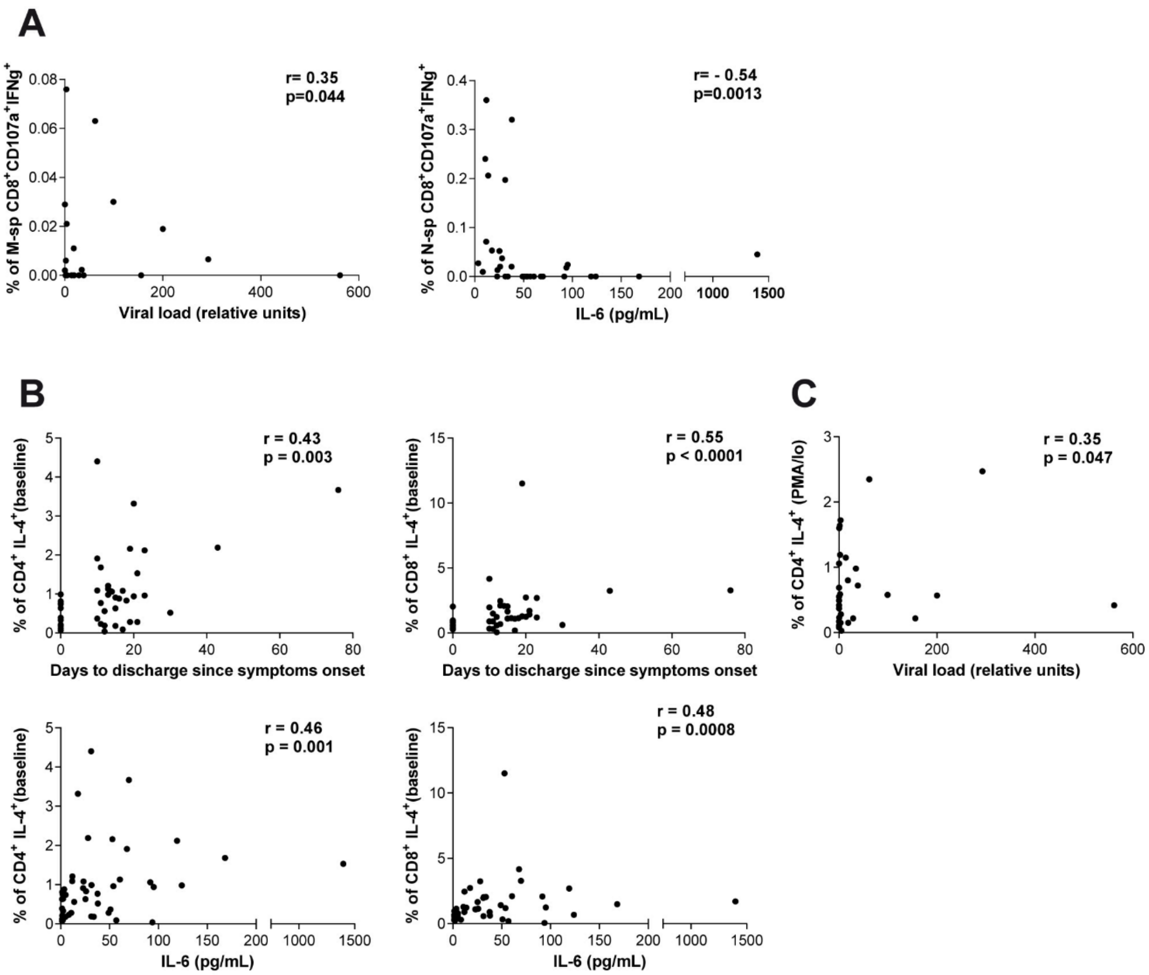


**Figure S3. Correlations between SARS-CoV-2-specific T cells and clinical parameters. (A)** Correlations between double CD107a$^+$IFNg$^+$ CD8$^+$T cells specific for the membrane (M) protein with viral load (left) and for the nucleocapsid (N) protein with IL-6 levels (right). Spearman rank correlation (n=33 for viral load and n=32 for IL-6). **(B)** Correlations between days to discharge since symptoms onset (top) or baseline IL-6 levels (bottom) and the frequency of CD4$^+$ (left) and CD8$^+$T cells(right) expressing IL-4 at baseline (unstimulated control). **(C)** Correlation between viral load and CD4$^+$T cells expressing IL-4 after PMA/Ionomycin. Spearman rank correlation (n=33 for viral load and n=46 for days to discharge since symptoms onset and IL-6).

### Figure S4

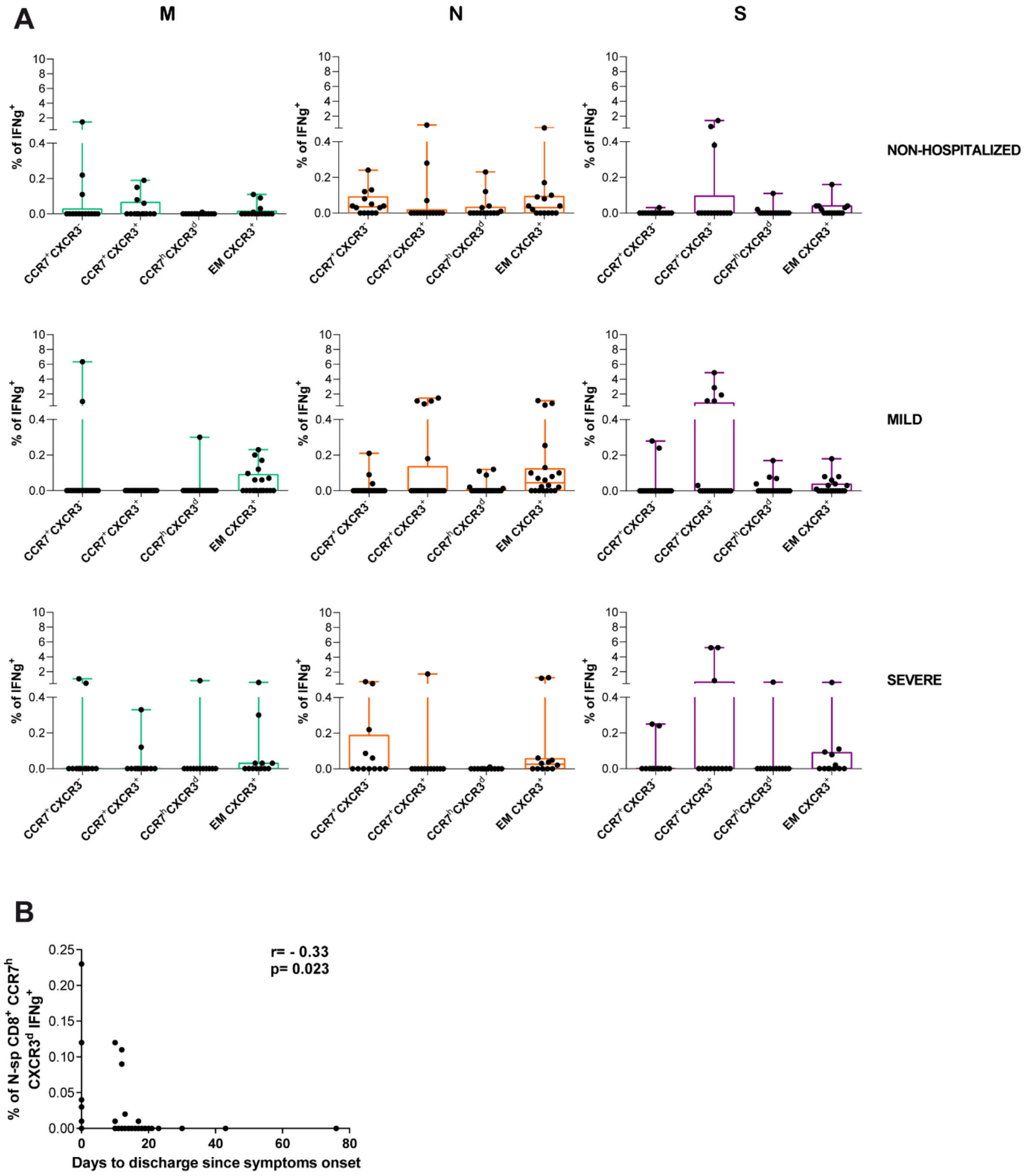


**Figure S4. IFNg expression in SARS-CoV-2-specific CD8$^+$ T cell subsets during acute infection. (A)** Net frequency of IFNg expression in CCR7$^+$CXCR3$^-$, CCR7$^+$CXCR3$^+$, CCR7$^h$CXCR3$^d$ and EM CXCR3$^+$ subsets within CD8$^+$ T cells after stimulation with any of the three viral proteins (membrane (M), nucleocapsid (N) and spike (S) proteins). Data are shown as median and upper range, where each dot represents an individual patient for each group (non-hospitalized n=14; mild n=20 and severe n=12). Statistical comparisons were performed using Kruskal-Wallis rank-sum test with Dunn's multiple comparison test. **(B)** Correlation between days to hospital discharge since symptoms onset and nucleocapsid-specific $CD8^+$ $CCR7^hCXCR3^d$ $IFNg^+$ T cells. Spearman rank correlation (n=46).

#### Figure S5

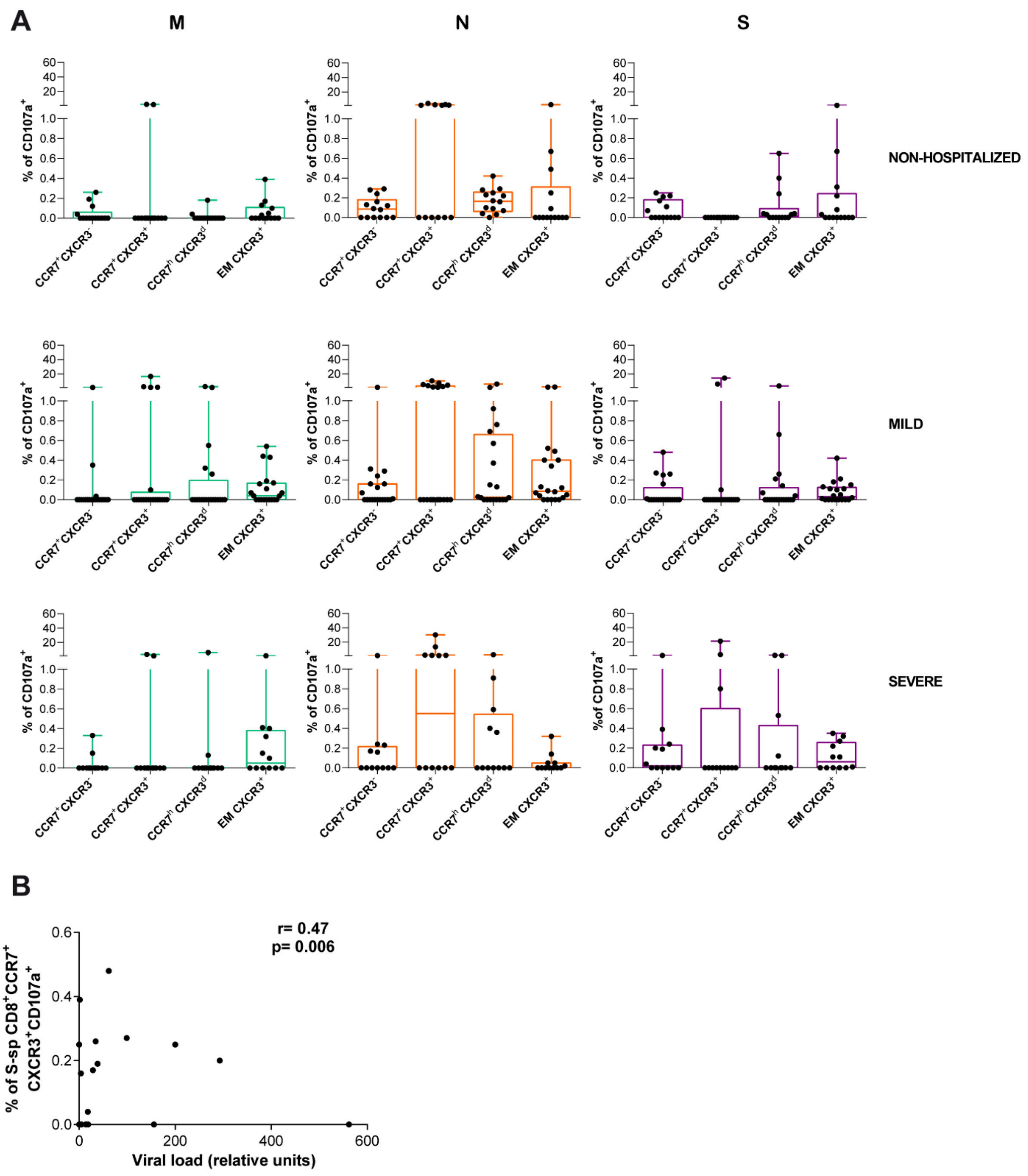

**Figure S5. CD107a expression in SARS-CoV-2-specific CD8$^+$ T cell subsets during acute infection.** **(A)** Net frequency of CD107a expression in CCR7$^+$CXCR3$^-$, CCR7$^+$CXCR3$^+$, CCR7$^h$ CXCR3$^d$ and EM CXCR3$^+$ subsets within CD8$^+$ T cells after stimulation with any of the three viral proteins (membrane (M), nucleocapsid (N) and spike (S) proteins). Data are shown as median and upper range, where each dot represents an individual patient for each group (non-hospitalized n=14; mild n=20 and severe n=12). Statistical comparisons were performed using Kruskal-Wallis rank-sum test with Dunn's multiple comparison test. **(B)** Correlation between viral load and the frequency of spike-specific CD8$^+$ CCR7$^+$CXCR3$^+$ CD107a$^+$ T cells. Spearman rank correlation (n=33).

### Figure S6

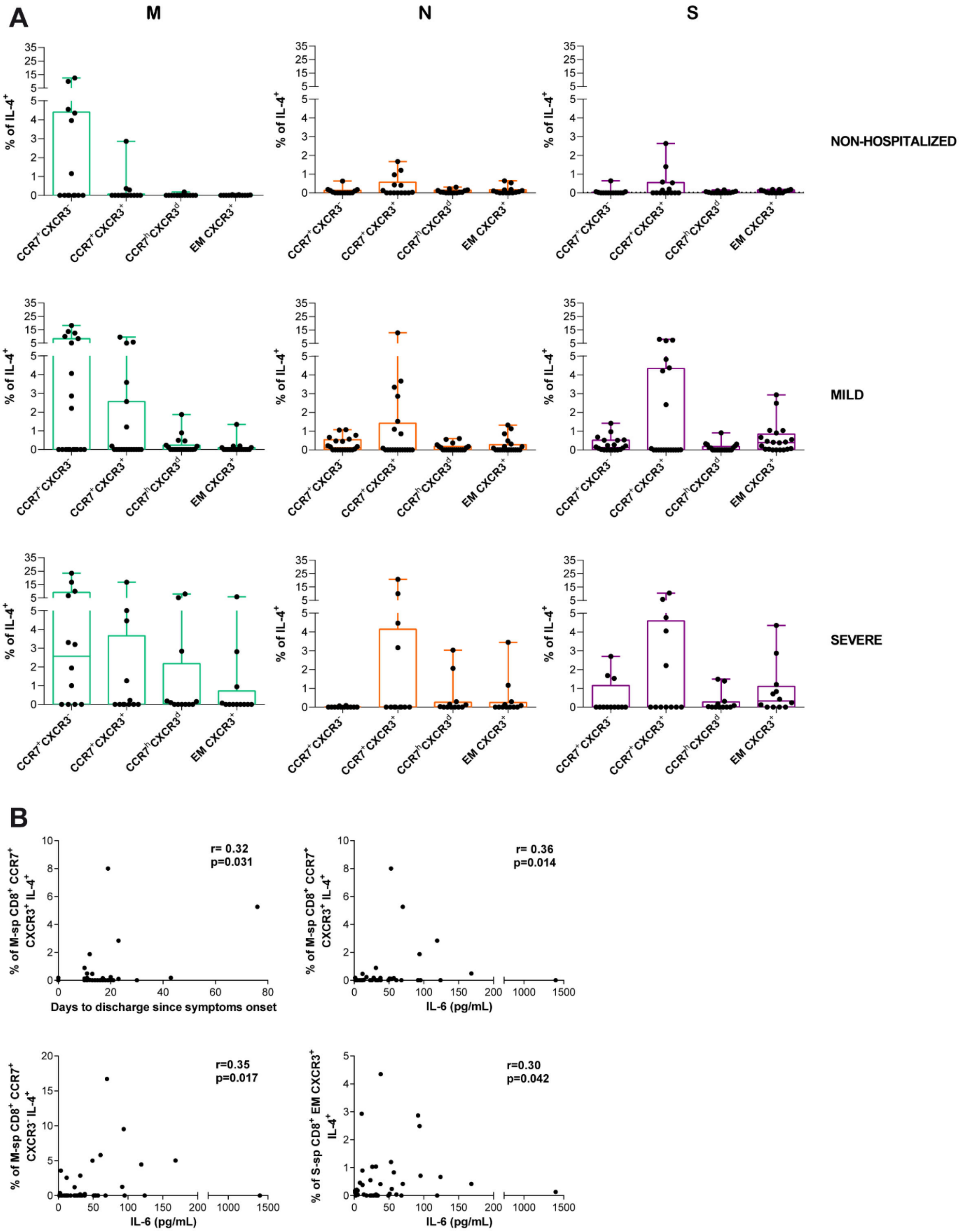

A
M
N
S
% of IL-4+
CCR7+CXCR3-
CCR7+CXCR3+
CCR7hiCXCR3d
EM CXCR3+
NON-HOSPITALIZED
MILD
SEVERE
B
r= 0.32
p=0.031
Days to discharge since symptoms onset
% of M-sp CD8+ CCR7+ CXCR3+ IL-4+
r= 0.36
p=0.014
IL-6 (pg/mL)
r=0.35
p=0.017
% of M-sp CD8+ CCR7+ CXCR3- IL-4+
r=0.30
p=0.042
% of S-sp CD8+ EM CXCR3+ IL-4+

**Figure S6. IL-4 expression in SARS-CoV-2-specific CD8$^+$ T cell subsets during acute infection.**

**(A)** Net frequency of IL-4 expression in CCR7$^+$CXCR3$^-$, CCR7$^+$CXCR3$^+$, CCR7$^h$CXCR3$^d$ and EM CXCR3$^+$ subsets within CD8$^+$ T cells after stimulation with any of the three viral proteins (membrane (M), nucleocapsid (N) and spike (S) proteins). Data are shown as median and upper range, where each dot represents an individual patient for each group (non-hospitalized n=14; mild n=20 and severe n=12). Statistical comparisons were performed using Kruskal-Wallis rank-sum test with Dunn's multiple comparison test. **(B)** Correlations between days to hospital discharge since symptoms onset or baseline IL-6 levels and the frequency of membrane or spike-specific CCR7/CXCR3 subsets of CD8$^+$ IL-4$^+$ T cells. Spearman rank correlation (n=46).

### Figure S7

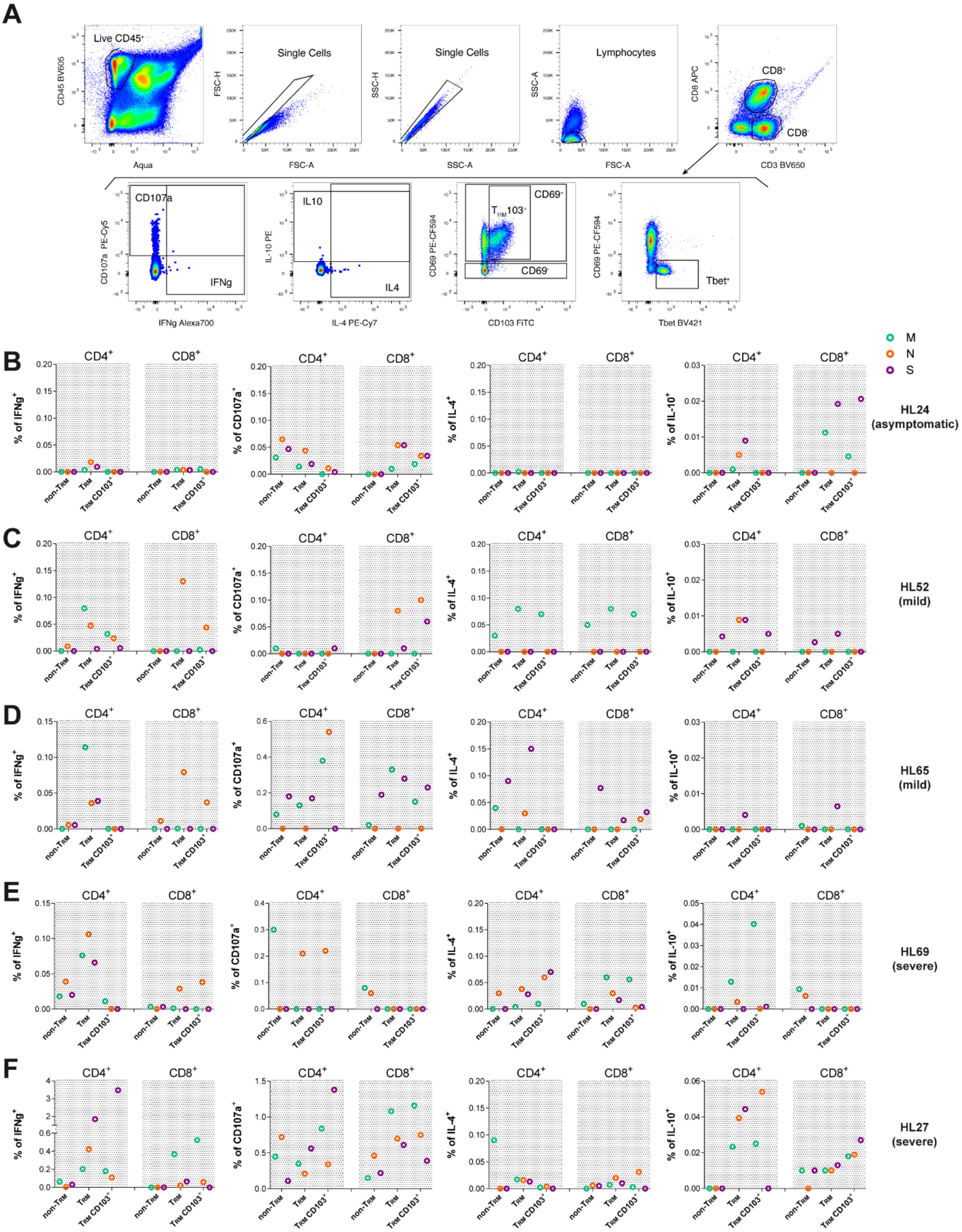

A
Live CD45+
Single Cells
Lymphocytes
CD8+
CD8-
CD107a
IFNg
IL10
IL4
CD69+
TRM103+
CD69-
Tbet+
B
C
D
E
F
M
N
S
HL24 (asymptomatic)
HL52 (mild)
HL65 (mild)
HL69 (severe)
HL27 (severe)
CD4+
CD8+
% of IFNg+
% of CD107a+
% of IL-4+
% of IL-10+
non-TRM
TRM
TRM CD103+

**Figure S7. Functional analysis of lung-resident SARS-CoV-2-specific T cells.**

**(A)** Representative flow cytometry plots for phenotypic analysis of lung-derived SARS-CoV-2-specific $CD4^+$ and $CD8^+$ T cells from patient HL27. Gating strategy consisted on a live $CD45^+$ gate, followed by doublet exclusion, a lymphocyte gate based on FSC vs. SSC and then a T cell gate based on $CD3^+$ $CD8^+$ or $CD8^-$ (putative $CD4^+$); from there we identified different functions (IFNg, CD107a, IL-4, IL-10) and resident phenotypes. Phenotype was based on the expression of CD69, CD103 and T-bet, where tissue-resident memory T cells ($T_{RM}$) were $CD69^+$, with a fraction of them expressing CD103, and non-$T_{RM}$ were $CD69^-$. As shown, T-bet was associated to the non-$T_{RM}$ fraction. **(B-F)** Net frequencies of IFNg, CD107a, IL-4 and IL-10 expression in SARS-CoV-2-specific $CD4^+$ or $CD8^+$ non-$T_{RM}$, $T_{RM}$ and $T_{RM}$ $CD103^+$ from patient **(B)** (HL24), **(C)** (HL52), **(D)** (HL65), **(E)** (HL69) and **(F)** (HL27). Viral proteins are shown in color green (membrane protein, M), orange (nucleocapsid protein, N) and purple (spike protein, S).

**Table S1. Patient characteristics at baseline.**

| | **Non-hospitalized** n=14 | **Mild** n=20 | **Severe** n=12 | *P-value between groups* |
|---|---|---|---|---|
| Age (years), range [a] | 25-42 | 38-64 | 51-69 | ***<0.0001*** |
| Females, n (%) [b] | 10 (71.4) | 11 (55) | 4 (33.3) | *0.556* |
| Fever, n (% yes) [b] | 9 (64.2.3) | 17 (85) | 8 (66.6) | *0.844* |
| Dyspnea, n (% yes) [b] | 2 (14.2) | 15 (75) | 7 (58.3) | *0.101* |
| SAFI, median [IQR] [a] | 475 [475-475] | 466.0 [461-471] | 447 [429.3-460] | ***<0.0001*** |
| SAFI 48h, median [IQR] [a] | 475 [475-475] | 454.5 [448.3-461] | 342.5.0 [283.5-396] | ***<0.0001*** |
| Days from symptoms to discharge, median | 0 [0-0] | 12.5 [11-15.7] | 21 [17.5-28.2] | ***<0.0001*** |
| Charlson Comorbidity Index, median [IQR] | 0 [0-0] | 1 [0-2] | 2 [1.2-2] | ***0.0002*** |
| **Immunology and Virology** | | | | |
| Leucocytes (10^9/L), median [IQR] [a] | 6.1 [5.4-7.3] | 5.7 [4.6-7.3] | 7.2 [4.9-9.3] | *0.462* |
| Lymphocytes (%), median [IQR] [a] | 33.5 [29.3-42.3] | 21.7 [15-25.5] | 15 [11.2-19.2] | ***<0.0001*** |
| Monocytes (%), median [IQR] [a] | 7 [6.5-8.2] | 8.2 [6.6-9.9] | 5.7 [2.9-9.5] | *0.243* |
| Neutrophils (%), median [IQR] [a] | 55.4 [49.2-61] | 69.75 [61.88- | 77.4 [64.6-79.75] | ***0.0006*** |
| Eosinophils (%), median [IQR] [a] | 2.6 [1.3-5.1] | 0.2 [0-0.7] | 0 [0-0.07] | ***<0.0001*** |
| Platelets (10^9/L), median [IQR] [a] | 291 [254.3-310.3] | 185.5 [158.3-297] | 191 [113.3-292.3] | *0.073* |
| Viral load (relative to housekeeping), median | 0.2 [0.1-4.9] | 3.5 [0.2-71.3] | 3.32 [0.8-165.6] | *0.215* |
| **Biochemistry** | | | | |
| D-dimer, (ng/ml), median [IQR] [a] | 94 [54.7-122] | 181 [130-401] | 339 [247-965] | ***0.0003*** |
| Ferritin, (ng/ml), median [IQR] [a] | 147 [30-202.0] | 336 [164.8-537.8] | 550.5 [309-1416] | ***0.0002*** |
| IL-6, (pg/ml), median [IQR] [a] | 1.7 [1.5-3] | 25.6 [12.3-28.2] | 55.3 [40.7-94.24] | ***<0.0001*** |

Statistical comparisons between groups (non-hospitalized, mild and severe) were performed using [a]Kruskal-Wallis rank-sum test with Dunn's multiple comparison test and/or [b]Chi-square test.

**Table S2. Spearman's correlation test between clinical parameters and the net frequency of SARS-CoV-2-specific CD4+ and CD8+ T cells by function.**

| | **Days to hospital discharge since symptoms onset (**n=46) | **IL-6 (pg/ml)** (n=46) | **SAFI ratio (48h)** (n=46) |
|---|---|---|---|
| **CD4+ T cells** | | | |
| **M peptides** | | | |
| IFN-γ | *p= 0.003*<br>r= 0.42 | *p= 0.0009*<br>r= 0.47 | *p= < 0.0001*<br>r= -0.5761 |
| CD107a | p= 0.107 (NS)<br>r= 0.24 | p= 0.505 (NS)<br>r= 0.10 | *p= 0.0335*<br>r= -0.3142 |
| IL-4 | p= 0.718 (NS)<br>r= 0.05 | p= 0.970 (NS)<br>r= -0.005 | p= 0.9834 (NS)<br>r= -0.003158 |
| IL-10 | *p= 0.028*<br>r= -0.32 | *p= 0.035*<br>r= -0.31 | *p= 0.0224*<br>r= 0.336 |
| **N peptides** | | | |
| IFN-γ | *p= 0.027*<br>r= 0.32 | *p= 0.099*<br>r= 0.24 | *p= 0.0015*<br>r= -0.4558 |
| CD107a | p= 0.929 (NS)<br>r= 0.013 | p= 0.252 (NS)<br>r= 0.17 | p= 0.0841 (NS)<br>r= -0.2575 |
| IL-4 | p= 0.627 (NS)<br>r= -0.07 | p= 0.595 (NS)<br>r= -0.08 | p= 0.8662 (NS)<br>r= 0.02554 |
| IL-10 | p= 0.122 (NS)<br>r= -0.23 | p= 0.144 (NS)<br>r= -0.21 | *p= 0.0222*<br>r= 0.3366 |
| **S peptides** | | | |
| IFN-γ | *p= 0.002*<br>r= 0.43 | *p= 0.007*<br>r= 0.39 | *p= < 0.0001*<br>r= -0.5585 |
| CD107a | p= 0.193 (NS)<br>r= 0.19 | p= 0.075 (NS)<br>r= 0.26 | *p= 0.0025*<br>r= -0.4356 |
| IL-4 | p= 0.273 (NS)<br>r= 0.16 | p= 0.391 (NS)<br>r= 0.12 | p= 0.2026 (NS)<br>r= -0.1914 |
| IL-10 | p= 0.760 (NS)<br>r= 0.04 | p=0.647 (NS)<br>r= 0.07 | p= 0.7723 (NS)<br>r= 0.04385 |
| **All peptides** | | | |
| IFN-γ | *p= 0.002*<br>r= 0.44 | *p= 0.001*<br>r= 0.45 | *p= < 0.0001*<br>r= -0.6165 |
| CD107a | p= 0.158 (NS)<br>r= 0.21 | *p= 0.024*<br>r= 0.33 | *p= 0.0002*<br>r= -0.5168 |
| IL-4 | p= 0.424 (NS)<br>r= 0.12 | p= 0.444 (NS)<br>r= 0.11 | p= 0.3377 (NS)<br>r= -0.1446 |
| IL-10 | p= 0.057 (NS)<br>r= -0.28 | p= 0.133 (NS)<br>r= -0.22 | *p= 0.0173*<br>r= 0.3494 |
| **CD8+ T cells** | | | |
| **M peptides** | | | |
| IFN-γ | p= 0.414 (NS)<br>r= 0.12 | p= 0.182 (NS)<br>r= 0.20 | p= 0.4156 (NS)<br>r= -0.123 |
| CD107a | p= 0.109 (NS)<br>r= 0.23 | *p= 0.003*<br>r= 0.41 | p= 0.1126 (NS)<br>r= -0.2371 |
| IL-4 | *p= 0.037*<br>r= 0.30 | p= 0.050 (NS)<br>r= 0.29 | *p= 0.0485*<br>r= -0.2925 |
| IL-10 | p= 0.612 (NS)<br>r= -0.07 | p= 0.429 (NS)<br>r= 0.11 | p= 0.5744 (NS)<br>r= 0.08499 |
| **N peptides** | | | |
| IFN-γ | p= 0.278 (NS)<br>r= 0.16 | p= 0.252 (NS)<br>r= -0.17 | p= 0.9502 (NS)<br>r= 0.009477 |
| CD107a | p= 0.254 (NS)<br>r= -0.17 | p= 0.226 (NS)<br>r= -0.18 | p= 0.0811 (NS)<br>r= 0.2599 |
| IL-4 | p= 0.761 (NS)<br>r= 0.04 | p= 0.800 (NS)<br>r= -0.03 | p= 0.5156 (NS)<br>r= -0.09832 |
| IL-10 | p= 0.431 (NS)<br>r= -0.11 | p= 0.273 (NS)<br>r= -0.16 | p= 0.4244 (NS)<br>r= 0.1207 |

| | | | |
|---|---|---|---|
| **S peptides** | | | |
| IFN-γ | p= 0.077 (NS)<br>r= 0.26 | p= 0.079 (NS)<br>r= 0.26 | p= 0.1173 (NS)<br>r= -0.2342 |
| CD107a | p= 0.393 (NS)<br>r= 0.12 | p= 0.927 (NS)<br>r= 0.01 | p= 0.5507 (NS)<br>r= 0.09029 |
| IL-4 | *p= 0.0006*<br>r= 0.48 | *p= 0.0002*<br>r= 0.51 | *p= < 0.0001*<br>r= -0.5793 |
| IL-10 | p= 0.552 (NS)<br>r= 0.09 | p= 0.407 (NS)<br>r= 0.12 | p= 0.6569 (NS)<br>r= -0.06726 |
| **All peptides** | | | |
| IFN-γ | *p= 0.015*<br>r= 0.35 | p= 0.535 (NS)<br>r= 0.09 | p= 0.0762 (NS)<br>r= -0.2641 |
| CD107a | p= 0.816 (NS)<br>r= -0.03 | p= 0.945 (NS)<br>r= 0.01 | p= 0.2961 (NS)<br>r= 0.1574 |
| IL-4 | p= 0.061 (NS)<br>r= 0.27 | *p= 0.042*<br>r= 0.29 | *p= 0.0158*<br>r= -0.3538 |
| IL-10 | p= 0.394 (NS)<br>r= -0.12 | p= 0.974 (NS)<br>r= 0.004 | p= 0.3317 (NS)<br>r= 0.1464 |

[a]SAFI ratio corresponds to the percentage of oxyhemoglobin saturation ($SaO_2$) in relation to the percentage of inspired oxygen ($FiO_2$) after 48h of hospitalization.
